# Blood flow restriction added to usual care exercise in patients with early weight bearing restrictions after cartilage or meniscus repair in the knee joint: A feasibility study

**DOI:** 10.1101/2022.03.31.22272398

**Authors:** Thomas Linding Jakobsen, Kristian Thorborg, Jakob Fisker, Thomas Kallemose, Thomas Bandholm

## Abstract

**Purpose:** In musculoskeletal rehabilitation, blood flow restriction – low load strength training (BFR-LLST) is theoretically indicated – as opposed to traditional heavy strength training – in patients who can or may not heavily load tissues healing from recent surgery. The main purpose was to examine the feasibility of BFR-LLST added to usual care exercise early after cartilage or meniscus repair in the knee joint.

**Methods:** We included 42 patients with cartilage (n=21) or meniscus repair (n=21) in the knee joint. They attended 9 weeks of BFR-LLST added to a usual care exercise at an outpatient rehabilitation center. Outcome measures were assessed at different time points from 4 (baseline) to 26 weeks postoperatively. They included: Adherence, harms, knee joint and thigh pain, perceived exertion, thigh circumference (muscle size proxy), isometric knee-extension strength, self-reported disability and quality of life.

**Results:** On average, patients with cartilage and meniscus repair performed >84 % of the total BFR-LLST supervised sessions. Thirty-eight patients reported 146 adverse events (e.g., dizziness) - none considered serious. A decrease in thigh circumference of the operated leg was not found in both groups from baseline to the end of the intervention period with no exacerbation of knee joint or quadriceps muscle pain.

**Conclusions:** BFR-LLST added to usual care exercise initiated early after cartilage or meniscus repair seems feasible and may prevent disuse thigh muscle atrophy during a period of weight bearing restrictions. Harms were reported, but no serious adverse events were found. Our findings are promising but need replication using RCT-design.

## Background

Cartilage or meniscus repair in the knee joint are common orthopedic procedures, in which patients are allowed early partial lower extremity weight bearing only. The consequence is a pronounced and persistent decrease of knee-extension strength in the operated leg (Stein et al. 2009; Van Assche et al. 2010; Hall et al. 2015; Lepley et al. 2015). The loss of muscle strength may delay return to normal daily activities (e.g., stair climbing), work/sports activities and negatively affect quality of life. A novel exercise modality to increase muscle strength is moderate blood flow restriction during low-load strength training (BFR-LLST) - also called occlusion training (Loenneke et al. 2011; Slysz et al. 2016). BFR-LLST involves application of a wrapping device such as an inflated tourniquet/cuff or an elastic band (Loenneke et al. 2011) to restrict the arterial inflow and venous outflow to muscle(s) during exercise. BFR-LLST (20-40% of 1-Repetition Maximum (RM)) requires much less external loading compared to traditional strength training (70-80% of 1RM). BFR-LLST produces positive training adaptations, such as muscle hypertrophy and increased strength in the lower extremity in healthy subjects, and patients with knee pathology (Loenneke et al. 2012; Slysz et al. 2016; Tennent et al. 2017; Hughes et al. 2017). The underlying mechanisms are not fully understood, but may stem from a complex interplay of reduction in oxygen delivery to the muscle (hypoxia), accumulation of metabolites, muscle fiber recruitment and proliferation of myogenic stem cells (Nielsen et al. 2012; Pearson and Hussain 2015; Scott et al. 2015). Few studies have shown promising results when patients, who are recovering from knee surgery, followed a rehabilitation program with BFR-LLST postoperatively (Takarada et al. 2000; Ohta et al. 2003; Tennent et al. 2017; Gaunder et al. 2017; Hughes et al. 2019b).

To our knowledge, early BFR-LLST added to usual care exercise has never been investigated in patients recovering from cartilage or meniscus repair in the knee joint, although some recommend BFR-LLST clinically to enhance the return of quadriceps function and facilitate an earlier return to sport (Sherman et al. 2020). In clinical practice, potentially limiting factors for the application and effect of BFR-LLST are acute knee joint and quadriceps muscle pain, perceptual responses, and fear of adverse events (Nakajima et al. 2006; Spranger et al. 2015; Hughes et al. 2019a; Cristina-Oliveira et al. 2020).

The aim of this study – investigating the feasibility of BFR-LLST added to usual care exercise early after cartilage or meniscus repair – was to examine whether 1) patients adhered to the BFR-LLST protocol, 2) patients experienced adverse events during the intervention period, 3) perceived exertion, quadriceps muscle pain and knee joint pain responses during BFR-LLST would change in a training session or over multiple training sessions, and 4) changes in thigh circumference over time. Additionally, clinical outcomes were assessed to describe changes over time when patients with cartilage and meniscus repair followed BFR-LLST added to usual care exercise.

## Material and Methods

This exploratory, prospective, and pragmatic study used consecutive sampling to assess the feasibility of 9 weeks of BFR-LLST added to a usual care exercise program at rehabilitation center, Section for Orthopaedic and Sports Rehabilitation (SOS-R), Nørrebro, City of Copenhagen. BFR-LLST knee-extensions was added to the usual care exercise program to enhance neuromuscular adaptations of the quadriceps muscle (Cook et al. 2018). BFR-LLST knee-extension without external load was added to the usual care exercise program the first 6 weeks postoperatively. At 6 weeks postoperatively, the BFR-LLST knee-extension with external load replaced the usual care knee-extension strength training exercise. Being an exploratory feasibility study, we designed the study with a flat outcome structure-having multiple evenly-valued outcome measures. Patients were assessed three times individually from week 4 to week 6 postoperatively. From the week 7 to week 12 postoperatively, patients were assessed 12 times during the 6-week bi-weekly group-based training sessions, and at 16 and 26 weeks postoperatively (See Figure 1).

**Figure 1.**
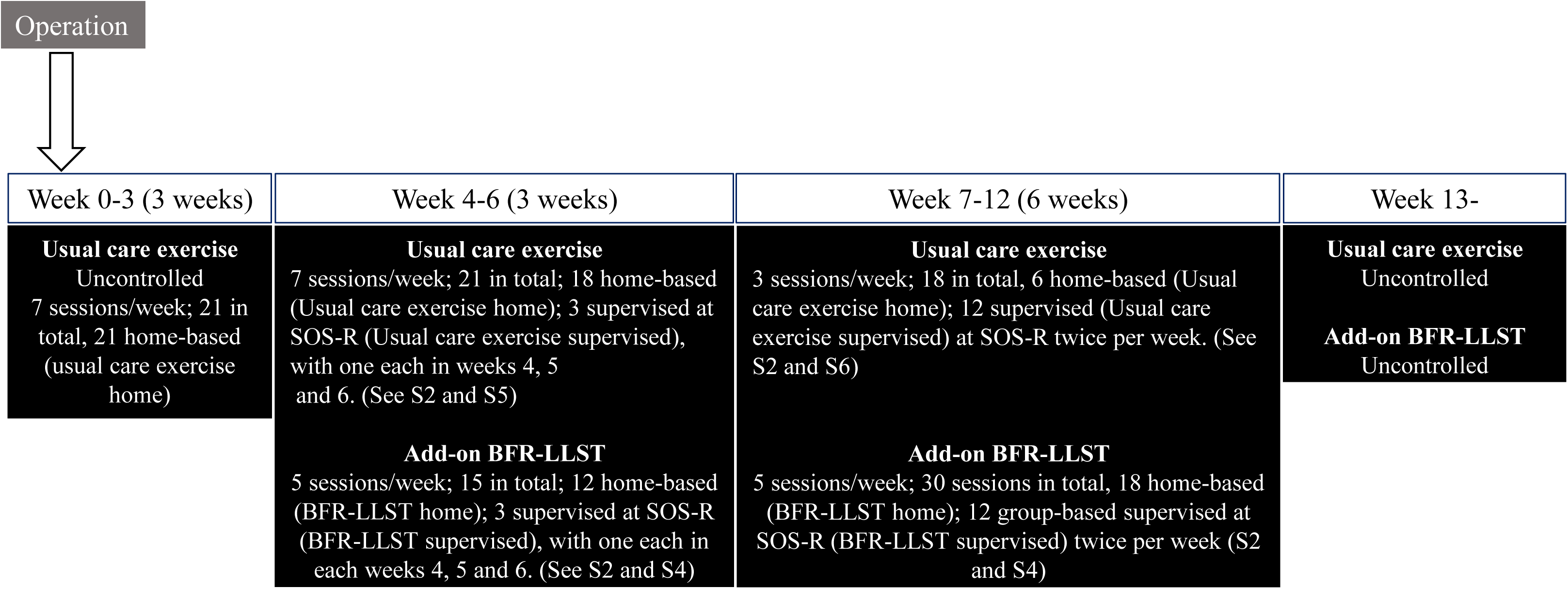
Overview of study treatment, testing and assessment timeline Abbreviations: Uncontrolled; Treatment provided prior to study inclusion or after the intervention period. SOS-R; Section for orthopaedic and sports rehabilitation. BFR-LLST; Blood flow restricted – low load strength training.

The reporting follows the CONSORT extension for randomized pilot and feasibility trials guidelines (Boutron et al. 2008; Eldridge et al. 2016), as well as the Consensus on Exercise Reporting Template (CERT) (Slade et al. 2016) (S1, S2).

Forty-two patients (cartilage (n=21) and meniscus (n=21) repair) were recruited postoperatively from a referral list at the SOS-R between December 13, 2017 and November 20, 2018. All patients were operated at two hospitals in the Copenhagen Area (Amager-Hvidovre and Bispebjerg-Frederiksberg). To be included, patients had to be between 18 and 70 years of age, have a cartilage or meniscus repair in 1 or 2 knee(s) and were not allowed full weight bearing (See S3). Patients were excluded if they were unable to speak or understand Danish/English or had problems that severely limited ambulatory function from unstable orthopedic (besides cartilage or meniscus repair in the knee joint). Additionally, we followed a combination of different risk assessment tools to avoid serious harms (Kacin et al. 2015; Patterson et al. 2019; Hughes et al. 2019b). As a result, patients with neurological, vascular or cardiac conditions; were pregnant, had cancer (current diagnosis), active infection; or a history of a) diagnosed major psychiatric disorder, b) illicit drug use, c) abusing alcohol or medication, d) endothelial dysfunction, peripheral vascular disease, hypertension, or diabetes, or e) heart disease and deep vein thrombosis, were excluded.

### Procedures

All patients had 4 different types of exercise during their scheduled 9 weeks of rehabilitation intervention period at SOS-R (Figure 1); 1) BFR-LLST supervised, which consisted of 3 individual and 12 group-based supervised sessions; 2) BFR-LLST home consisted of 30 home-based sessions; 3) Usual care (non-BFR) exercise supervised was 3 individual and 12 group-based supervised sessions; and 4) Usual care (non-BFR) exercise home consisted of 24 home-based sessions. After the scheduled 9 weeks of rehabilitation intervention period, the physical therapists responsible for supervised usual care exercise decided whether the patients should continue their rehabilitation.

At each supervised session of BFR-LLST added to usual care exercise (15 sessions in total) from week 4 to week 12 postoperatively, the following acute outcome measures were assessed: adherence, adverse events related to the BFR-LLST, external load during BFR-LLST, knee joint and quadriceps muscle pain before (at rest), during (4 sets each) and after (at rest) the BFR-LLST, perceived exertion during BFR-LLST (4 sets each), and thigh muscle size indicated by circumference for the operated and healthy leg. Additionally, the maximum knee joint and quadriceps muscle pain during the usual care exercise supervised session was assessed by patients’ recall immediately after the session. Additional clinical outcome measures, such as active and passive knee joint range of motion, knee joint effusion, self-reported function and knee-related quality of life, and self-reported functional ability to complete specific activities were recorded at baseline (first individually assessment), and at 16 and 26 weeks postoperatively. At 26 weeks postoperatively, isometric knee extension and flexion muscle strength were assessed. Outcome assessors were experienced physical therapist and they were not blinded to the treatment provided. Most of the additional assessments was conducted by the principal investigator (TLJ).

As this study was pragmatic, we were unable to fully control if the patients received exactly 9 weeks of BFR-LLST added to usual care exercise as listed in the trial registry. Eleven weeks are considered more likely on average, because 1) patients were referred and allowed to perform BFR-LLST earlier than expected, and 2) there was often a time delay from last individual BFR-LLST supervised (6 weeks postoperatively) to the first group-based BFR-LLST supervised (approximately 7 weeks postoperatively) session.

Some between-hospital variation in the content of the rehabilitation regimes existed, but generally the added BFR-LLST knee-extension exercise adhered to the following restrictions for patients with cartilage and meniscus repair. BFR-LLST knee-extensions from 90° of knee flexion to full knee extension with no external load was allowed 1 and 2 weeks postoperatively. At 6 weeks, BFR-LLST knee-extensions from 90° of knee flexion to full knee extension was allowed with external load. Full weight bearing was tolerated from 0 to 90 degrees knee joint flexion from week 7 postoperatively (Detailed Rehabilitation regimes, See S3).

### Exercise intervention

#### Blood flow restriction – low-load strength training (BFR-LLST)

The BFR-LLST protocol was the same for both patients with cartilage or meniscus repair and outlined in Table 1 (Toigo and Boutellier 2006; Kronborg et al. 2014; Scott et al. 2015). At baseline, the physical therapist determined the individual patient’s limb occlusion pressure (LOP), which is defined as the minimum occlusion pressure necessary required to stop the flow of arterial blood into the lower limb distal to a pneumatic cuff (Noordin et al. 2009). This procedure was used to calculate the relative LOP (percentage of LOP) to enhance the treatment effect, while limiting perceived discomfort and risk of adverse events during BFR-LLST (Murray et al. 2020). The LOP in the lower limb was identified by increasing the pressure in a 20-cm wide pneumatic cuff with a sphygmomanometer (Heine Gamma® G5, HEINE, Optotechnik GmbH & Co., Herrsching, Germany) placed on the most proximal portion of the thigh until pulse stop, while the patients were sitting on an examination couch and the heel resting on a chair or the floor with the knee joint angle between 45 and 90 degrees of flexion attached. Pulse stop was registered on a finger clip connected to a portable valid and reliable hand-held oximeter attached to the patients’ second toe (Brekke et al. 2020). To account for variability associated with the determination of the LOP, the individual patient’s LOP was subtracted by 20 mmHg. From this adjusted LOP, the relative applied 80% LOP was calculated. When exercising at very low intensity (20% of 1RM), 80% LOP has been proposed to increase hypertrophy (Lixandrão et al. 2015) and muscle activity (Fatela et al. 2016) in the quadriceps muscle. After individualizing the pressure, the patients were thoroughly instructed in how to perform unilateral BFR-LLST for the knee-extension exercise with a cuff (same as used for LOP determination) at the SOS-R, and at home with an elastic band (Trithon Knee Wraps, Trithon Sport, Denmark) (S2, S4). The first 6 weeks postoperatively, patients performed BFR-LLST without external loads. At 6 weeks postoperatively, BFR-LLST was performed using weight band(s) fixated around the ankle. At each BFR-LLST supervised session, the external load was increased by 0.5 to 1.0 kilograms, if the patients could perform more than 15 repetitions in the 4^th^ and last set. The patients were instructed to perform BFR-LLST at home in the same manner with the elastic band tightened proximal to the thigh and lifting similar external loads, using weight band(s) provided by the SOS-R.

**Table 1.** The BFR-LLST protocol for the knee-extension exercise

| Variables | Knee-extension exercise |
| --- | --- |
| Load, repetition maximum | 30 (~20-40 % of 1RM) |
| Repetitions per set | ≤ 6 weeks: 30, 15, 15, max. 30*<br>> 6 weeks: 30, 15, 15, 15 |
| Sets per session | 4 |
| Rest between sets, seconds | 30-45 |
| Sessions per week | 5 |
| Duration of the experimental period, weeks | Approximately 9 weeks; 3 weeks without and 6 weeks with external loads |
| Contraction modes, seconds | 1 isometric, 2 eccentric, 2 concentric |
| Rest between repetitions, seconds | 0 |
| Time under tension, seconds | ≤ 6 weeks: 450 (5 seconds x (30+15+15+30 reps))<br>> 6 weeks: 375 (5 seconds x (30+15+15+15 reps)) |
| Contraction failure in each set | No, only the last set out of 4* |
| Range of motion, degrees | Max. 90 |
| Rest between training sessions, hours | 24 to 48 hours |
| Anatomical definition of the exercise (exercise form) | Yes |
| Mode per week | ≤ 6 weeks: 3 times supervised in total. 4-5 times at home.<br>> 6 weeks: 2 times supervised, 3 times at home. |
| Cuff application | As proximal as possible on the lower limb |
| Cuff type (materials) | 20-cm width (Supervised with cuff; Home with elastic band). |
| Occlusion pressure | Cuff: 80% limb occlusion pressure (LOP). Elastic band: Should be as tight as the cuff corresponding to 80% LOP. |
| BFR-LLST time (total) per session, seconds | ≤ 6 weeks: 585 (9 ¾ min.) (450 time under tension + 3x45 rest)<br>> 6 weeks: 510 (8 ½ min.) (375 time under tension + 3x45 rest) |
| Duration of the entire BFR-LLST session | Max. 10 minutes twice per week (supervised) and max. 10 minutes 3 times per week at home. |

\*Patients started the BFR-LLST without external load the first 6 weeks postoperatively due to weight-bearing restrictions and the number of repetitions were limited to 30 in the 4<sup>th</sup> and last set.

#### Usual care exercise

At the first treatment (baseline) approximately 3 weeks postoperatively, patients were instructed in a usual care exercise program, which consisted of 4 strengthening and 2 range of motion exercises according to the rehabilitation regimes for cartilage and meniscus repair (S2, S5). Exercises were to be performed daily at home. Additionally, at baseline the physical therapist instructed patients in correct gait patterns and how they should manage and control their knee joint pain and swelling. At 6 weeks postoperatively, the usual care exercise was replaced with a new criteria-based usual care exercise program that patients performed at home and at SOS-R (S2, S6). The 60-minute new usual care exercise program consisted of pre-warm-up (10 minutes); warming-up exercises (10 minutes); progressive strength training exercises targeting ankle plantar flexors, quadriceps, hamstring, and hip abductor and adductors muscles; balance exercises and flexibility exercises (30-35 minutes). The unilateral knee-extension progressive strength training exercise to increase quadriceps strength was replaced by the BFR-LLST knee-extension to limit mechanical strain on and protect the operated knee joint (Sherman et al. 2020). We recommended patients to perform BBF-LLST knee-extension (10-15 minutes) after the usual care exercise program. The traditional progressive strength training exercises were performed to volitional muscular failure with the following descriptors: 3 sets of 12 repetitions using an intensity of 12 RM and time under tension of 4 seconds (2 seconds concentric and eccentric contractions, respectively) (Toigo and Boutellier 2006). When the patients mastered the strength training exercise in a specific muscle group correctly with acceptable patient-perceived knee symptoms/pain, the exercise was replaced with a more demanding weight-bearing strength training exercise within the same muscle group in the usual care exercise program. The patients received the program in hard-copy and in a digital-version (Exorlive, Oslo, Norway) sent via email (S6). We were unable to control the rehabilitation that patients received at the hospital prior to study inclusion. Generally, they were instructed to perform basic unloaded strengthening and range of motion exercises daily comparable to the program received at the first treatment (baseline) in this study.

### Clinical application (adherence)

At each BFR-LLST added to the usual care exercise session, number of training sessions performed for BFR-LLST (supervised and home), usual care exercise (supervised and at home) and the BFR-LLST descriptors (LOP applied, number of sets, repetitions and external load lifted) were recorded. BFR-LLST descriptors at home were patient-reported via a training diary (S4). Furthermore, the specific exercises performed at each usual care exercise supervised session were noted.

### Harms

At each visit, any adverse events potentially related to BFR-LLST at SOS-R or at home were reported by the patients to the physical therapist responsible for each BFR-LLST supervised session. Patients were interviewed based on a pre-defined and standardized questionnaire of potential adverse events related to BFR-LLST (dizziness, quadriceps muscle pain at rest, knee joint pain at rest, bruising, numbness in the lower leg, subcutaneous bleeding (bruising), cardiovascular or respiratory complaints, deep venous thrombosis, other events or no events) (Nakajima et al. 2006). If a patient experienced clinical signs and symptoms of deep venous thrombosis at home (Mazzolai et al. 2018), they were urged to contact a medical doctor or the physical therapist at SOS-R immediately (S4). An adverse event was considered serious if it caused death, was life-threatening, or resulted in persistent or significant disability/incapacity or required inpatient hospitalization according to European Medicines Agency (European Medicines Agency 2006). A serious event would require a permanent discontinuation of the BFR-LLST intervention. Additionally, all adverse events or complications were documented regardless of its perceived relation to the exercise intervention, operation or occurrences not related to the study. The number of possible adverse events were summed.

### Outcome measures

Acute outcome measures were recorded at each BFR-LLST supervised session. Thigh muscle size was indicated by a small line 15 cm proximal to the base of patella and measured by using a standard tape measure circumference of the thigh (Järvelä et al. 2002; Ezaki et al. 2010; Jakobsen et al. 2010; Tennent et al. 2017). The value was recorded to the nearest 0.1 cm. Moreover, knee joint and quadriceps muscle pain was assessed using a 0–100-mm visual analog scale (VAS-mm) with end points of “no pain” and “worst pain imaginable” (Breivik et al. 2008), and maximum rating of perceived exertion was measured using the Borg scale ranging from 6 (no exertion at all) to 20 (maximal exertion) by the patients’ recall immediately after each BFR-LLST set (Borg 1970). At baseline, 16- and 26-week assessments, additional outcome measures were recorded. They were: knee joint range of motion measured with a large (30 cm long-armed) universal goniometer (Jakobsen et al. 2010), and knee joint effusion using a standard tape measure placed 1 cm proximal to the base of patella (Jakobsen et al. 2010). Knee self-reported function and quality of life (Roos et al. 1998) was assessed using the Knee Injury and Osteoarthritis Outcome Score (KOOS; including subscales of symptoms, pain, activities of daily living, function in sport/recreation, and knee-related quality of life) with scores ranging from 0–100 (Roos et al. 1998). To address self-reported functional status, we used the patient-specific functional scale (PSFS), where the patients scored, on a scale ranging from 0-10, their ability to complete self-selected important activities they were unable or had difficulty in performing (Stratford et al. 1995). The average score of maximum 5 activities was calculated. Both self-reported questionnaires were scored on a worst to best scale. At 26 weeks postoperatively, maximal isometric knee-extension and -flexion strength in 60 degrees of knee flexion was measured using a handheld dynamometer (MicroFet 2, Hoogan Scientific, Salt Lake City, US) fixated between the patients’ distal tibia and the resistance pad of a leg extension/leg curl strength training machine. The resistance pad was fixated by the load of the weight stacks of the training machine and could not be moved. (Gagnon et al. 2005; Mikkelsen et al. 2016). Patients had one practice trial followed by minimum 3 and maximum 10 knee-extensions with verbal encouragement. The test was ended if the isometric knee extension strength decreased in 2 consecutive trials or the maximum of 10 trials was reached (Aalund et al. 2013). The highest strength value was used at the data point. The same procedure was used to determine maximal isometric knee flexion strength.

### Sample size

No formal sample size calculation was performed due to the descriptive character of the study and no efficacy testing was performed (Arain et al. 2010). Approaches to the sample size justification for feasibility studies vary greatly (Billingham et al. 2013). It has been recommended to target a sample size between 15 (Julious 2005) and 50 (Sim and Lewis 2012) for pilot/feasibility studies. We targeted a sample size of 40 patients with a full outcome dataset and continued recruitment until this was achieved (Figure 2). We aimed for an equal distribution of patients with cartilage (n=20) and meniscus (n=20) repair. We considered a sample of 40 patients large enough to indicate acceptability of the intervention, evaluation of the study protocol (for a potential future large-scale trial), and to provide enough data for future population-specific sample size estimations (Kaur et al. 2017). To consider for dropouts during the study, we included 51 patients. To allow for some attrition after the last patient in each group was included, we enrolled 42 patients in total.

**Figure 2.**
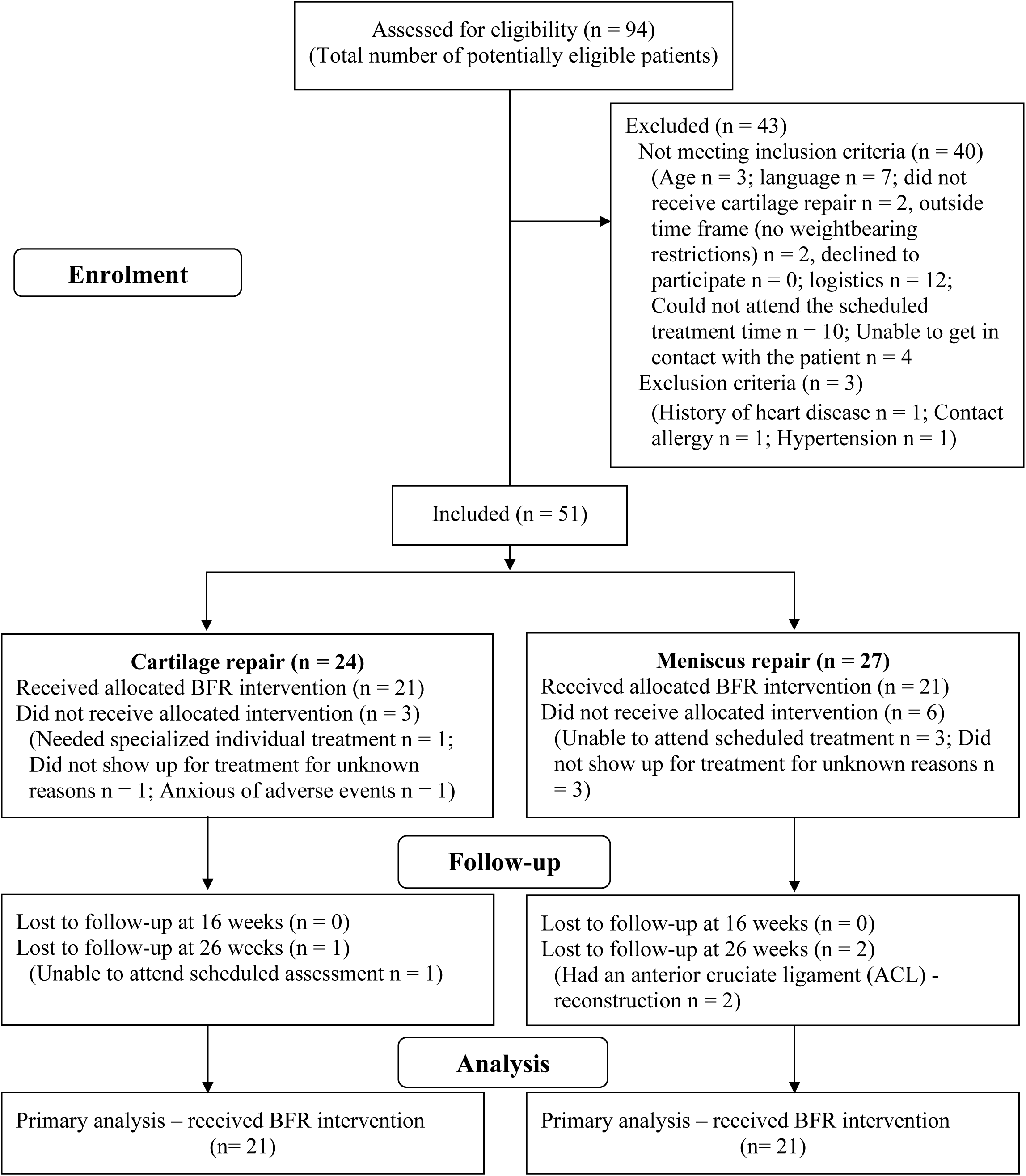
Patient flow diagram

### Statistical analysis

We used a linear mixed-effect model to analyze 1) within-group changes for all outcome measures over time from baseline to the different time points (assessments during each BFR-LLST added to the usual care exercise supervised session, 16- and 26-week postoperatively) and 2) within-set changes from the 1^st^ to 4^th^ set (4 sets each) recorded during each BFR-LLST supervised session (15 sessions per patient) in all patients. A type 3 test for the linear mixed-effect model was used to evaluate the overall effect from the 1^st^ to 4^th^ set during the BFR-LLST supervised session. Changes in knee joint and quadriceps pain at rest from before to after each BFR-LLST supervised session, and the difference in maximal knee joint pain during usual care exercise and the BFR-LLST supervised sessions, were examined using a Wilcoxon signed-rank test. No multiple comparisons were performed for any individual timepoints. Assumption of normal distribution was evaluated using Q-Q plots, histograms and Shapiro-Wilk tests. The goodness of model fit to the data was evaluated for normality of residuals and variance homogeneity by residual vs. predicted outcome plots. The completed training sessions performed in percentage was calculated by summarizing the performed number of training sessions divided by the total number of training sessions scheduled. Missing values in the linear mixed effect models were considered ignorable under the assumption of data being missing at random (MAR) and the likelihood estimation used in the models (Little and Rubin 2014).

All data were entered in EpiData Entry, version 3.3 (Epidata, Odense, Denmark). Analyses were performed using R version 1.4.1103 (R Foundation for Statistical Computing, Vienna, Austria), Microsoft Excel programs (Microsoft Office 365, 2019, Redmond, WA, US) and Graphpad Prism version 8.3.1 (GraphPad Software, San Diego, California, US). A P-value less than 0.05 was considered statistically significant.

## Results

Ninety-four patients with cartilage or meniscus repair were assessed for eligibility from December 2017 to March 2019 (Figure 2), which was the total number of potentially eligible patients during that time period. Of those 94 patients, 43 patients were excluded. Fifty-one patients were included. Four patients withdrew from the study for reasons unrelated to the BFR-LLST. Four patients did not show up for their treatment, and we were unable to get in contact with them by phone or email. Nothing in their data (complaints, adverse events, complications) indicated that the reason for dropping out, was related to the BFR-LLST. One patient dropped out before the BFR-LLST started due to anxiety of potential adverse events. In total, 42 patients with cartilage (n=21) and meniscus (n=21) repair received the BFR-LLST intervention, and characteristics of the patients at baseline, and 16- and 26-week assessments are presented in Table 2 and S7 Table 1.

**Table 2.** Baseline characteristics for the patients with cartilage and meniscus repair

| Characteristics | Cartilage (n=21) | Meniscus (n=21) |
| --- | --- | --- |
| Age, mean ± 1SD, years | In their 30s* | In their 30s* |
| Women/men, no (%) | 5 (24)/16 (76) | 8 (38)/13 (62) |
| Body mass index, mean ± 1SD, kg/m<sup>2</sup> | 26.1 ± 3.6 | 24.9 ± 3.5 |
| Smoker, no (%) | 4 (19) | 5 (24) |
| Repair type, (no) | Patella (5)/FT (8)/MFC (6)/LFC (2) | LM (5)/MM (15)/LM+MM (1) |
| Cartilage size damage<sup>a</sup>, median (IQR), cm<sup>2</sup> | 2.30 (1.45 to 3.25) |  |
| Multi-operations<sup>b</sup>, no (%) | 12 (57) | 11 (52) |
| Right/left operated knee, no (%) | 9 (43)/12 (57) | 15 (71)/6 (29) |
| Same knee operated before, no (%) | 10 (48) | 7 (33) |
| Opposite knee operated before, no (%) | 6 (29) | 4 (19) |
| Postoperative complications<sup>c</sup>, no (%) | 2 (10) | 3 (14) |
| Supervised rehabilitation prior to baseline assessment, no (%) | 5 (24) | 3 (14) |
| Tried BFR before baseline assessment, no (%) | 5 (24) | 5 (24) |
| Accustomed to exercise, no (%) | 18 (86) | 18 (86) |
| Have used pain medication before baseline assessment, no (%) | 5 (24) | 2 (10) |
| Acetaminophen (Paracetamol), no (%) | 5 (24) | 2 (10) |
| NSAID, no (%) | 4 (19) | 2 (10) |
| Opioids and opioid-like drugs, no (%) | 2 (10) | 1 (5) |
| Hospital performing the operation, AHH/BBH, no (%) | 8 (38)/13 (62) | 5 (24)/16 (76) |
| Time from operation to baseline assessment, median (IQR), weeks | 2.0 (1.9 to 5.0) | 2.3 (2.0 to 2.6) |

SD; Standard Deviation. IQR; Interquartile Range. LM; Lateral meniscus. MM; Medial meniscus. FT; Femur throchlea. MFC; Medial femur condyle. LFC; Lateral femur condyle; BFR; Blood flow restriction. NSAID; Non Steroidal Anti-Inflammatory Drug. AHH; Amager-Hvidovre Hospital. BBH; Bispebjerg Hospital. <sup>a</sup>n=20. <sup>b</sup>ACL-reconstruction (n=6), <sup>c</sup>Knee stiffness, muscle tightness, knee pain, quadriceps hematoma.
*\*All data on age were collected at the first visit to the rehabilitation center. Wording chosen to comply with medRxiv's policy regarding limiting the number of indirect identifiers.*

### Clinical application (adherence) and training characteristics

On average, patients with cartilage or meniscus repair completed more than 80% of the BFR-LLST supervised, BFR-LLST at home, usual care exercise supervised and usual care exercise at home (S7 Table 2). On average, the BFR-LLST added to the usual care exercise intervention period lasted 11 (SD±1.2) weeks. The individualized applied limb occlusion pressure (LOP) varied from 104 to 176 mmHg during the BFR-LLST supervised, and on average the patients performed the recommended 75 repetitions per BFR-LLST supervised session approximately 79% of the time (S7 Table 2). The median external load during the BFR-LLST knee-extension exercise increased from 0 kg at baseline to 2.5 kg at the end of the BFR-LLST supervised intervention period (median = 0.28 kg/week, range = 0.23-0.32 kg/week, p < 0.001) (S7 Table 2, S7 Table 3), but overall remained low (median = 1 kg, range = 0-7 kg) (S7 Table 2). The number and name of the exercises performed during the group-based usual care exercise supervised program at SOS-R are shown in S7 Table 4.

### Outcome measures

#### Knee joint and quadriceps muscle pain

Using the classification by the International Association for the Study of Pain (IASP), patients experienced none to mild (Treede et al. 2019) maximal knee joint pain during BFR-LLST supervised (median VAS-mm = 0, IQR = 0-12), which was significantly and clinically meaningful (Farrar et al. 2001; Dworkin et al. 2008) lower than the maximal knee joint pain experienced during usual care exercise supervised (median VAS-mm =23, IQR 0-40) (p < 0.001) (S7 Table 2). On the contrary, patients experienced moderate to severe maximum quadriceps muscle pain during BFR-LLST supervised (median VAS-mm = 47, IQR 12-70), which was significantly and clinically meaningful (Dworkin et al. 2008) higher than the maximal quadriceps muscle pain during usual care exercise supervised (median VAS-mm = 0, IQR 0 to 6) (p < 0.001). The median value of the knee joint and quadriceps muscle pain at rest before and after BFR-LLST supervised session was zero (all median VAS-mm = 0) (S7 Table 2).

#### Changes during the BFR-LLST intervention period

During the 11-week BFR-LLST intervention period, the thigh circumference of the operated leg, increased, on average, 0.12 cm per week in the meniscus repair group (p < 0.001), and a similar pattern (although not statistically significant) was found in the cartilage repair group (0.05 cm per week, p = 0.099) (Figure 3A, S7 Table 3). Taken the thigh circumference of the healthy leg into account, the thigh circumference difference between legs diminished over time in the cartilage (-0.06 cm per week, p < 0.006) and meniscus group (-0.10 cm per week, p < 0.001) (S7 Table 3). Quadriceps muscle pain did not change over time in all patients (p = 0.330), while knee joint pain decreased over time in the meniscus repair group only (-1.1 VAS-mm per week, p < 0.010) (Figure 3B, S7 Table 3). Patients found the BFR-LLST to be slightly more demanding at the end compared to the beginning of intervention period, as the rating of perceived exertion (Borg Scale) increased with approximately 0.1 and 0.2 point per week for the cartilage repair group (p < 0.034) and meniscus repair group, respectively (p < 0.001) (Figure 3C, S7 Table 3).

**Figure 3.**
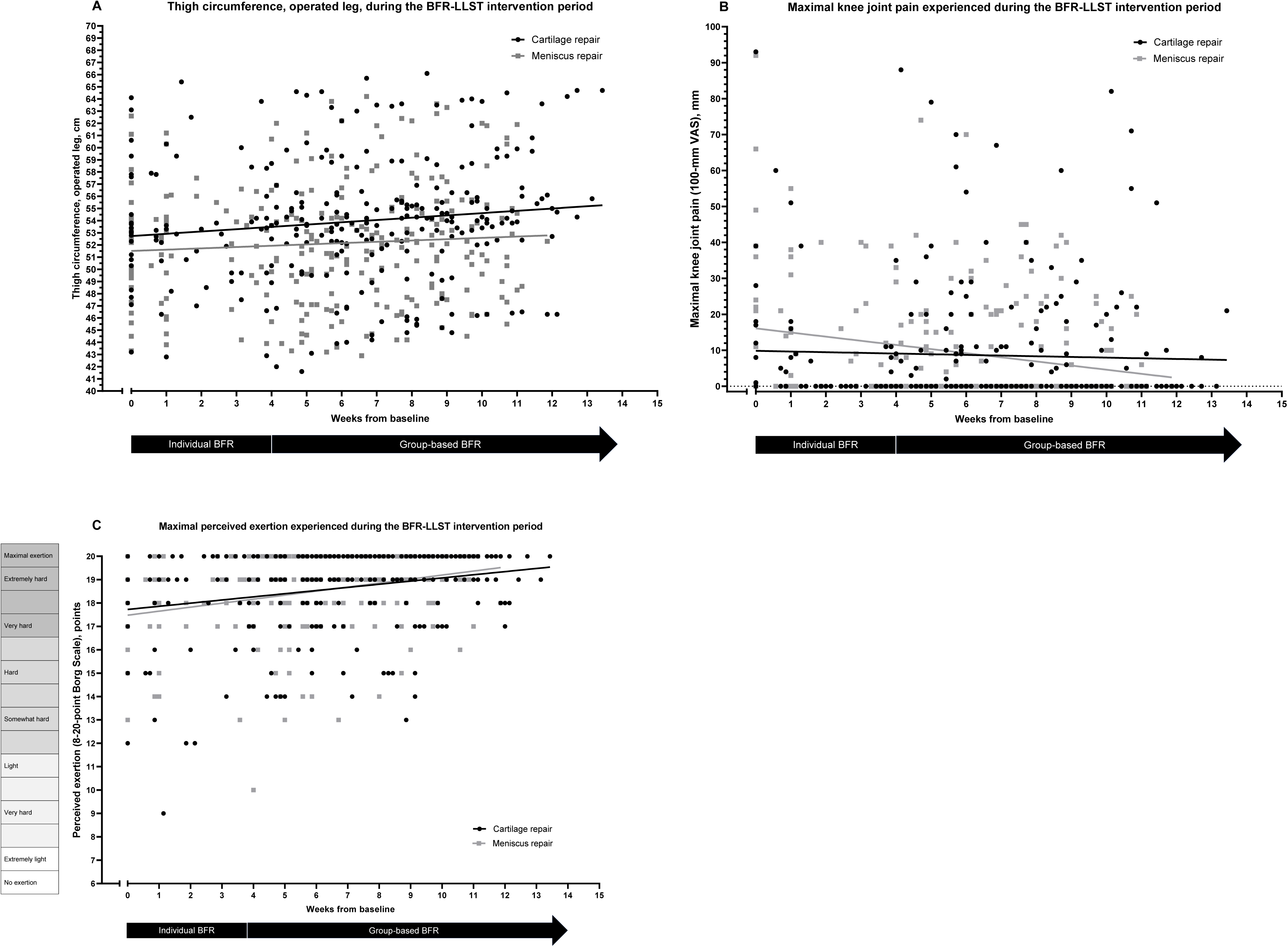
Scores of each patient as well for mean scores for the patients with cartilage (black dots and line) or meniscus repair (gray dots and line) over time for thigh circumference (A), knee joint pain (B) and perceived exertion (C) during the BFR-LLST intervention. BFR-LLST; Blood flow restriction – low load strength training. VAS; Visual Analog Scale.

#### Changes within the BFR-LLST supervised session (4 sets each)

On average, patient experienced none to mild (4.8 to 5.9 VAS-mm) knee joint pain and no difference between the 1^st^ to the 4^th^ set of BFR-LLST (p = 0.224) (Figure 4A, S7 Table 6). On the contrary, increases were found from the 1^st^ to the 4^th^ set for quadriceps muscle pain (Figure 4B, S7 Table 6) and rating of perceived exertion (Borg Scale) (Figure 4C, S7 Table 6) from 11.1 to 40.7 VAS-mm (p < 0.0001) and from10.8 to 18.5 points (p < 0.0001), respectively.

**Figure 4.**
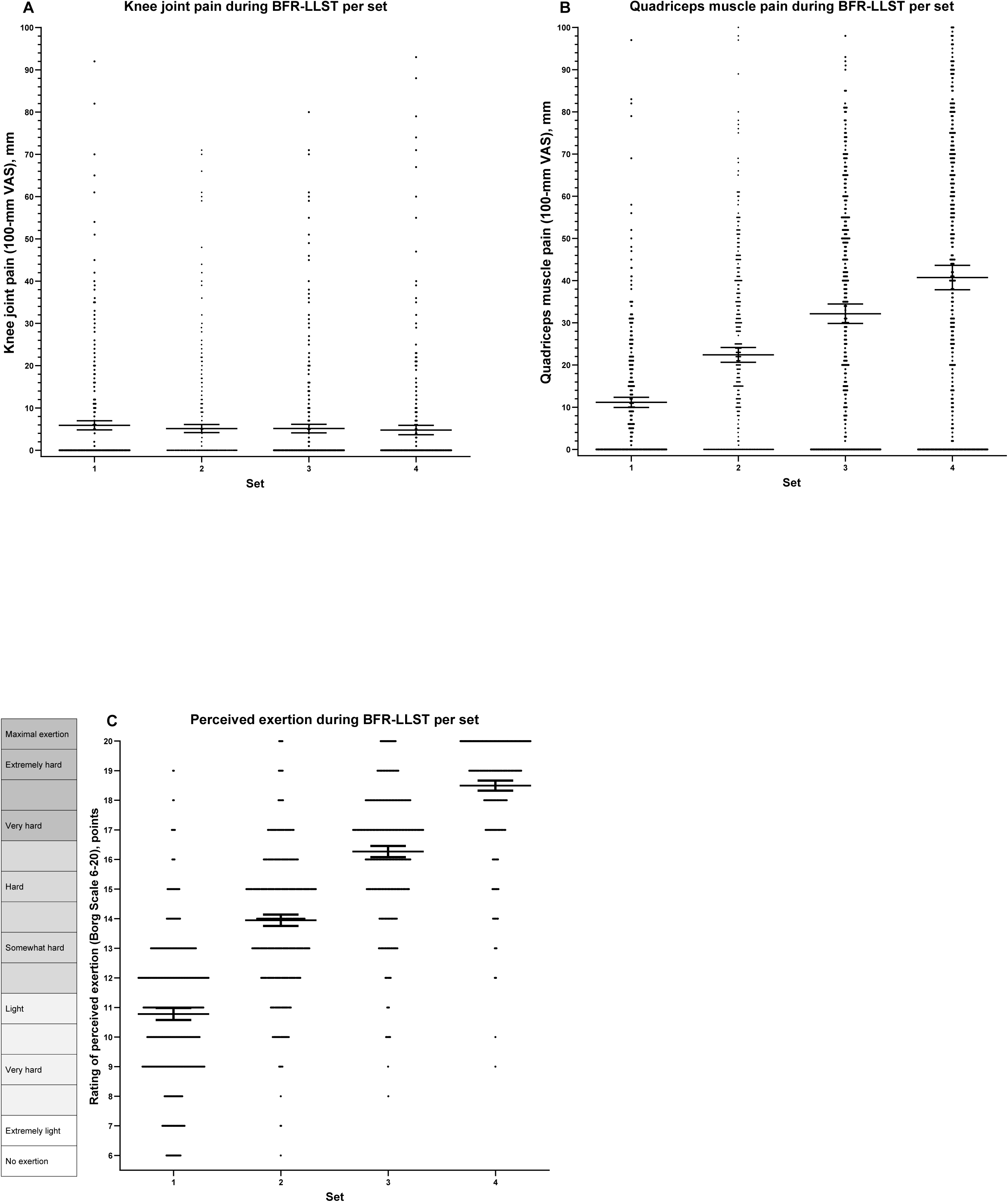
Knee joint pain (A), quadriceps muscle pain (B) and perceived exertion (C) from 1st to 4th set within the BFR-LLST supervised session (15 sessions each) for all patients. Scatterplots with means including whiskers representing 95% confidence intervals.

#### Clinical outcome measures

Changes of the clinical outcome measures from baseline to 16 and 26 weeks postoperatively in patients with cartilage and meniscus repair are presented in table S7 Table 7. Predominately, significant improvements were revealed from baseline to 16 or 26 weeks postoperatively for thigh circumference (operated leg), KOOS subscales, PSFS and active/passive knee joint range of motion of extension/flexion. No significant increase in knee joint effusion in the operated leg was seen from baseline to 26 weeks. At 26 weeks postoperatively, an isometric knee-extension muscle strength deficit of the operated compared to the healthy leg (Limb symmetry index (LSI) = 76 - 89%) existed. This isometric muscle strength deficit was small for knee-flexion (LSI= 94 - 96%) (S7 Table 7).

#### Harms

Thirty-eight out of 41 patients reported a total of 146 adverse events, but none were considered serious (S7 Table 5). Dizziness (52 cases) occurred for approximately half of the patients (n=19) - but no one fainted. All the adverse events were considered transient, meaning the symptoms disappeared within a short time frame after the completion of the BFR-LLST session, and no patients dropped out because of BFR-LLST.

At the 26 weeks postoperative assessment, 5 out of 21 (25%) and 8 of 19 (42%) of patients with cartilage and meniscus repair, respectively, reported postoperative complications (S7 Table 1). Knee joint pain was the most prevalent reported complication, which seemed related to the initiation of weight-bearing or impact activities.

## Discussion

This study investigated the feasibility of BFR-LLST added to usual care exercise in patients with early weight-bearing restrictions after cartilage or meniscus repair. The main findings were 1) patients adhered well to the BFR-LLST protocol, 2) the majority of patients reported adverse events – none considered serious, 3) during the BFR-LLST supervised session quadriceps muscle pain increased, while knee joint pain was consistently none to mild. At rest, knee joint and quadriceps muscle pain were none to mild during the BFR-LLST intervention period and 4) the thigh circumference (muscle mass proxy) in the operated leg did not decrease early after cartilage or meniscus repair.

### Interpretation

#### Clinical application (adherence) and training characteristics

Patients were able to adhere to the high-volume (5 times a week) high-intensity (80% LOP) BFR-LLST protocol, using both a pneumatic cuff during BFR-LLST supervised and a simple elastic band at home (BFR-LLST home). It should be noted that two patients had the relative LOP reduced either transient or permanently due to discomfort. As a result, adjustment of the relative LOP must be considered in patients experiencing a uncomfortable tight pneumatic cuff placed proximal to the thigh, even though little differences in discomfort during BFRT elbow flexion with relative LOPs ranging from 40% to 90% has been reported in healthy individuals (Loenneke et al. 2016). Only one in nine dropouts in the study was related to BFR-LLST. The patient was anxious about potential harms of BFR-LLST and dropped out prior to the first BFR-LLST supervised session at baseline.

#### Harms

Reporting of adverse events in a standardized and systematic way in randomized controlled trials examining exercise therapy (Niemeijer et al. 2020) or BFR exercise (Hughes et al. 2017) is rare. The focus is typically efficacy of new interventions, such as BFR exercise (Ioannidis et al. 2004). A systematic approach in registering adverse events seems imperative as serious and non-serious adverse events and concerns related to BFR exercise have been reported in practitioner- or individual-reported surveys (Nakajima et al. 2006; Patterson and Brandner 2018). Therefore, we conducted a prospective registered study examining patients with weight-bearing restrictions that focused on, and registered, adverse events related to BFR-LLST at each supervised session. Like other studies (Tennent et al. 2017; Hughes et al. 2019b), we did not find any serious adverse events (European Medicines Agency 2006), although the study was very likely underpowered to detect rare serious adverse events. However, we noted a higher frequency of transient dizziness compared to other clinical trials (Nakajima et al. 2006; Tennent et al. 2017; Hughes et al. 2019b) but similar to the experience of practitioners of BFR-LLST (Patterson and Brandner 2018). Half of the patients experienced dizziness while loosening the cuff after the BFR-LLST, which may be associated with hypotension or a vaso-vagal response (Patterson and Brandner 2018). We registered very few events of bruising or subcutaneous hemorrhage compared to a large survey on healthy individuals performing BFR exercise (Nakajima et al. 2006). Importantly, patients experienced more frequently itching in the lower leg, calf tightness and discoloration of the leg distal to the cuff. Finally, at the 26-week assessment, the patient-reported postoperative complications were surprisingly high in patients with meniscus repair, which has shown to be common and not related to the rehabilitation protocols (Nepple et al. 2012).

#### Outcome measures

##### Knee joint pain, quadriceps muscle pain and perceived exertion

Clinicians and orthopedic surgeons have concerns about knee joint pain and intra-articular damage during postoperative rehabilitation (Sherman et al. 2020). The maximum of knee joint pain experienced during the BFR-LLST was close to none, and lower than during usual care exercise, which promotes BFR-LLST for patients with cartilage or meniscus repair that require prolonged weight-bearing restrictions (Kruithof et al. 2018; Sherman et al. 2020). The reason for the lower pain levels could be the lower external load on the knee joint during BBFR-LLST knee-extension and/or the BFR-LLST-induced hypoalgesia effect (Korakakis et al. 2018). The maximum quadriceps muscle pain experienced during BFR-LLST was moderate and higher compared to close to none during usual care exercise, which is in line with patients performing BFR-LLST early after ACL-reconstruction (Hughes et al. 2019a). The perceived pain increase was temporary during the occlusion period, as knee joint and quadriceps muscle pain at rest before and after the BFR-LLST session was close to none, indicative of no exacerbating of pain symptoms over time.

As seen in healthy subject performing BFR exercise (Yasuda et al. 2011; Dankel et al. 2019), the BFR-LLST becomes gradually more demanding from the 1^st^ to the 4^th^ set, where patients on average experienced moderate quadriceps muscle pain and rated the perceived exertion as very/extremely hard in the last and 4^th^ set. The gradually reduction of blood flow to the quadriceps muscle during BBF-LLST impacts tissue metabolism and fatigue (Takarada et al. 2000; Hollander et al. 2010), which may explain the perceived quadriceps muscle pain (Fukuba et al. 2007) and exertion (Hollander et al. 2010). Interestingly, many patients in the present study did not register quadriceps muscle pain as much as increased exertion during the last and 4^th^ BFR-LLST set. This contradiction may be explained by multiple facts. Some patients associated quadriceps muscle pain during BFR-LLST (exercise-related) as a) beneficial to enhance recovery (Dannecker and Koltyn 2014), or b) had a high level of self-efficacy for tolerating muscle pain during exercise (Motl et al. 2007), and thereby did not perceive BFR-LLST as painful.

Patients experienced the same amount or slightly higher perceived exertion during the BFR-LLST over the entire intervention period, indicating that the BFR-LLST stimulus was constant as the external training load (kilo lifted) during BFR-LLST knee-extension increased. Additionally, quadriceps muscle and knee joint pain remained stable or tended to decrease over the intervention period. Similar patterns were found in patients with ACL-reconstruction over an 8-week BFR intervention period, which suggests that BFR-LLST might have a hypoalgesic effect on knee joint pain (Hughes et al. 2019a). The mechanism behind the reduction of knee joint pain during BFR-LLST is unclear, but the moderate quadriceps muscle pain originated from the ischemic and exercise-induced and/or the limb occlusion pressure during BFR-LLST may have had a pain modulating effect in the present study (Korakakis et al. 2018).

##### Thigh circumference, muscle strength and additional clinical outcomes

Thigh circumference (and the difference in thigh circumference between both legs) as surrogate proxy for thigh muscle mass did not decrease during the intervention period, even though patients followed an early range of motion and weight-bearing restrictions protocol after cartilage and meniscus repair. This is in line with research showing that BFR-LLST increase, or diminish disuse, muscle atrophy in patients after knee surgery (Takarada et al. 2000; Tennent et al. 2017) and healthy individuals (Slysz et al. 2016; Lixandrao et al. 2018; Centner et al. 2019). To our knowledge, no study has indicated the beneficial effect of BFR-LLST (or any other intervention) on disuse atrophy performed early after cartilage or meniscal repair without exacerbation of knee joint and quadriceps muscle pain. Still, we found a knee-extension muscle strength deficit of the operated compared to the non-operated leg (LSI) 26 weeks postoperatively in patients with cartilage and meniscus repair. Several reasons may explain the knee-extension deficit after knee surgery. First, the scheduled BFR-LLST dose with 4 sets at each BFR-LLST session, 5 times a week for 9 weeks were maybe not sufficient to counteract the negative disuse effects in the early postoperative phase with limited mobilization and weight-bearing. Second, the traditional knee-extension progressive high-load strength training was not part of the late rehabilitation program (at 6 weeks postoperatively) and this exercise may have increased the knee-extension strength more compared to BBF-LLST knee-extension (Hughes et al. 2017), but potentially increasing the number of postoperative complications (Steadman et al. 2003; Sherman et al. 2020). Third, complications after cartilage (Weber C. et al. 2018) and meniscus (Lind et al. 2013; Blanchard et al. 2020) repair are common and limiting the maximum effort patients may have put in to the knee-extension strength testing.

Overall, our KOOS scores were slightly better (5 to 15 points higher on KOOS pain, KOOS symptoms and KOOS ADL), and our knee-extension strength showed similar values (S7 Table 7), than results derived from prospective studies investigating either cartilage repair (Van Assche et al. 2010; Saris et al. 2014) or meniscus repair with (Lepley et al. 2015) or without (Pihl et al. 2021) ACL reconstruction surgery, between 6 to 2 years postoperatively. However, KOOS scores, especially the subgroups KOOS Sport/recreation and KOOS Quality of life, remained up to 25-30 points lower 6 months postoperatively than compared to a reference population (Paradowski et al. 2006). Our clinical outcomes (KOOS and lower limb strength) call for or require a long-term rehabilitation strategy after cartilage (Van Assche et al. 2010) and meniscus repair (Pihl et al. 2021).

### Study limitations

Several limitations should be noted. First, the study was a unblinded prospective feasibility study without a control group. Therefore, we cannot speak as to the efficacy of early BFR-LLST after cartilage and meniscus repair. A larger single- (or double-) blinded randomized controlled trial should be undertaken to test the hypothesis that BFR-LLST added to usual care exercise is superior to usual care without BFR-LLST, for knee-extension strength and physical function after cartilage or meniscus repair. Second, no objective registration was carried out to measure patients’ adherence to the BFR-LLST protocol at home. However, we stressed the importance of adhering to the BFR-LLST home program and patients were encouraged to full-fill their patient-reported training diary at each visit, which we believe resulted in the high reported adherence rate. Third, we had no pre-surgery recordings, and were unable to control the rehabilitation prior to the study inclusion at the rehabilitation center. After inclusion, patients followed a standardized rehabilitation program. Fourth, the results of this study may not be extrapolated to other BFR-LLST protocols. However, our BFR-LLST protocol was similar to the recommended BFR-LLST protocols to enhance muscle mass and strength (Scott et al. 2015; Patterson et al. 2019; Lorenz et al. 2021).

### Clinical applications (generalizability)

The study was a pragmatic, prospective feasibility study, where patients followed a usual care exercise pathway in a clinical practice setting, except BFR-LLST knee-extension without load was added to the program from week 4 to 6 week postoperatively and replaced the traditional progressive strength training with BFR-LLST knee-extension with external load from week 7 to 12 postoperatively. If a clinician follows the exclusion criteria and assess the patient’s complaint(s) at each supervised BFR-LLST session, we believe the results of this study can be transferred directly into clinical practice, as the study was conducted in a clinical setting, for patients with early weight-bearing restrictions after cartilage and meniscus repair in the knee joint. It should be noted that patients were primary young and had free access to their treatment.

## Conclusion

Nine weeks of BFR-LLST added to usual care exercise initiated early after cartilage and meniscus repair in the knee joint seems feasible without exabation of knee joint symptoms. Patients adhered well to the BFR-LLST protocol. Harms were reported (e.g. dizziness), but none were considered serious. No disuse atrophy of the thigh muscles was found, albeit patients had weight-bearing restrictions in the intervention period. The encouraging results call for a RCT to investigate the efficacy of early BFR-LLST in patients with similar or a higher level of disability.

## Data Availability

All data produced in the present study are available upon reasonable request to the authors

## List of abbreviations

BFR-LLST: Blood flow restriction – low load strength training
RCT: Randomized Controlled Trial
RM: Repetition Maximum
SOS-R: Section for Orthopaedic and Sports Rehabilitation
CERT: Consensus on Exercise Reporting Template
LOP: Limb Occlusion Pressure
VAS: Visual Analog Scale
KOOS: Knee Injury and Osteoarthritis Outcome Score
PSFS: Patient-Specific Functional Scale
MAR: Missing At Random
ACL: Anterior Cruciate Ligament
SD: Standard Deviation
IQR: Interquartile Range
LM: Lateral meniscus
MM: Medial meniscus
FT: Femur throchlea
MFC: Medial femur condyle
LFC: Lateral femur condyle
NSAID: Non-Steroidal Anti-Inflammatory Drug
IASP: International Association for the Study of Pain
LSI: Limb symmetry index

## Supporting information

**S1 Checklist CONSORT extension to randomized pilot and feasibility trials**

**S2 CERT (Consensus on Exercise Reporting Template)**

**S3 Rehabilitation regimens - Cartilage (Steadman procedure) and meniscus repair**

**S4 Blood Flow Restriction Exercise Leaflet**

**S5 Usual care exercise after cartilage and meniscus repair in the knee joint - week 3-6 postoperatively**

**S6 Usual care exercise after cartilage and meniscus repair in the knee joint - week 7 postoperatively**

**S7 Supplementary results**

## Declarations

### Ethics approval and consent to participate

The Committees on Biomedical Research Ethics for the Capital Region of Denmark approved the study (Protocol nr. 17010473), and the study was pre-registered at Clinical.Trials.gov (NCT03371901) December 13, 2017. Patients’ consent to retrieve personal medical data were sought, and therefore approval from the Danish Data Protection Agency was not necessary according to the Danish Data Protection Legislation and Danish Health Act. Patients were provided with written information about the purpose, procedures, and safety issues. Written informed consent was obtained in strict accordance with the Declaration of Helsinki.

### Consent for publication

Written informed consent from the individual, appearing in the Supporting information S4 Blood Flow Restriction Exercise Leaflet and S5 Usual care exercise after cartilage and meniscus repair in the knee joint - week 3-6 postoperatively Exercise Description, was obtained.

### Availability of data and material

The datasets used and/or analyzed during the current study are available from the corresponding author on reasonable request.

### Competing interests

The authors have declared that no competing interests exist, except TB, who declares: I have received speaker’s honoraria for talks or expert testimony on the efficacy of exercise therapy to enhance recovery after surgery at meetings or symposia held by biomedical companies (Zimmer Biomet and Novartis). I have received fees for writing textbook chapters (Munksgaard) and for organising post-graduate education, such as post-graduate courses in clinical exercise physiology (Danish Physical Therapy Organization) or PhD courses on clinical research methodology (University of Copenhagen). I am an editorial board member with Br J Sports Med. I am an exercise physiologist and physical therapist and may have a cognitive exercise bias.

### Funding

This study was supported by a grant from Praksisfonden (grant number R65-A1601). We certify that no party having a direct interest in the results of the research supporting this article has or will confer a benefit on us or on any organization with which we are associated.

### Authors’ contributions

All authors contributed to the conceptualization and methodology of the study, interpretation of data, wrote the work, drafting the article or revising it critically for important intellectual content and accepted final approval of the version to be published. Project administration, investigation, acquisition of data: TLJ, JF. Formal analyses: TLJ, TK.

## Acknowledgements

We thank physical therapists Mads Thorup Langelund, Christina Ramos Stavngaard, Andreas Olsen, Jonas Larsen and Mikkel Hvidsteen for data acquisition and valuable contribution to the standardization of the rehabilitation programs. We acknowledge Christian Vedel Sørensen for his help in analyzing and coding data. A sincere acknowledgement goes to the management and physical therapists working at the Centre of Rehabilitation, Nørrebro, City of Copenhagen, Denmark. Finally, we highly appreciate the financial support from Praksisfonden.

## S1 Checklist CONSORT extension to randomized pilot and feasibility trials

**CONSORT 2010 checklist of information to include when reporting a pilot or feasibility randomized trial in a journal or conference abstract**

| Item | Description | Reported on page number |
| --- | --- | --- |
| Title | Identification of study as randomised pilot or feasibility trial | 1 |
| Authors * | Contact details for the corresponding author | 1 |
| Trial design | Description of pilot trial design (eg, parallel, cluster) | 6 |
| Methods |  | 6-16 |
| Participants | Eligibility criteria for participants and the settings where the pilot trial was conducted | 6-7 |
| Interventions | Interventions intended for each group | 6-12, Figure 1, S2, S3, S4, S5, S6. |
| Objective | Specific objectives of the pilot trial | 6 |
| Outcome | Prespecified assessment or measurement to address the pilot trial objectives** | 7-8<br>13-14 |
| Randomization | How participants were allocated to interventions | N/A |
| Blinding (masking) | Whether or not participants, care givers, and those assessing the outcomes were blinded to group assignment | 8 |
| Results |  | 16-22 |
| Numbers randomized | Number of participants screened and randomised to each group for the pilot trial objectives** | 17 |
| Recruitment | Trial status† | 16 |
| Numbers analysed | Number of participants analysed in each group for the pilot objectives** | 16-17 |
| Outcome | Results for the pilot objectives, including any expressions of uncertainty** | 19-22, Figure 3 and 4.<br>S7 Supplementary results |
| Harms | Important adverse events or side effects | 22, S7 Table 5 |
| Conclusions | General interpretation of the results of pilot trial and their implications for the future definitive trial | 28 |
| Trial registration | Registration number for pilot trial and name of trial register | 29 |
| Funding | Source of funding for pilot trial | 30 |

Citation: Eldridge SM, Chan CL, Campbell MJ, Bond CM, Hopewell S, Thabane L, et al. CONSORT 2010 statement: extension to randomised pilot and feasibility trials. *BMJ*. 2016;355.
*\*this item is specific to conference abstracts*
*\*\*Space permitting, list all pilot trial objectives and give the results for each. Otherwise, report those that are a priori agreed as the most important to the decision to proceed with the future definitive RCT.*
*†For conference abstracts.*

## S2 CERT (Consensus on Exercise Reporting Template)

### 1. Type of exercise equipment

Initially, all patients received a written description of the BFR-LLST protocol explaining the advantages, disadvantages and potential risks related to the BFR-LLST (S4 Blood Flow Restriction Exercise Leaflet) intervention. Furthermore, the patients received hand-outs with two different exercise programs for i) the immobilization period week 3-6 (S5), and ii) group session period 7-12 (S6), respectively. The exercise template used during group-based usual care exercise supervised sessions were also used for the home training in the same period of as well as the following period from week 13. All exercises were illustrated with pictures and a thorough written description (S6).

List of training equipment needed during the BFR-LLST added to the usual care exercise program:

- 20-cm wide pneumatic cuff with a sphygmomanometer attached, 41-59 cm circumference, Heine Gamma® G5, HEINE, Optotechnik GmbH & Co., Herrsching, Germany.
- An elastic band (Trithon Knee Wraps, Trithon Sport, Denmark) for BFR-LLST home-based.
- Weight band(s) fixated around the ankle with loads of 0.5, 1, 1.5, 2, 3, 4, 5, 6, 7 kilograms
- A spinning bike
- A Concept 2^®^ rowing machine
- Carpet tiles
- Variation of dumbbells or kettlebells x 2 to match the exact strength level of the patient
- Barbell with plates matching the strength level of the patient
- Ball (diameter ≈22 cm)
- Body bar
- Theraband^®^ elastic band sheet in green or red colour
- Reebok^®^ step box
- Seated leg press machine (Cybex® VR3, Medway, MA, USA)
- BOSU^®^ - balance trainer (Diameter 65 cm)
- Wobblesmart^®^ balance board

### 2. Instructor qualifications

All physical therapists had a minimum two years of experience with rehabilitation of patients with cartilage and meniscus repair. The physical therapists were familiar with BFR-LLST for patients with weight-bearing restriction and/or knee joint pain. To ensure standardization of the BFR-LLST added to the usual care exercise program, the primary investigator and the clinical responsible physical therapist held a 2-hour instruction session with all physical therapists that carried out the group-based usual care exercise supervised sessions.

### 3. Individual or group

Patients had three individual visits during the first six weeks postoperatively. From approximately week seven postoperatively, patients attended at bi-weekly group-based BFR-LLST added usual care exercise supervised program (15 sessions).

### 4. Supervision

Handouts of all exercise programs and instructions were handed to the patients at the study site. The patients performed the exercises at the Section for Orthopaedic and Sports Rehabilitation (SOS-R) Copenhagen as well as at home.

Twelve group-based sessions were planned for the six-week period (BFR-LLST added to usual care supervised). The group-based sessions included up to eight patients per class. The sessions lasted 60 minutes each and were held twice per week with one or two days in between. The patients started participating in the group-based BFR-LLST added to usual care exercise supervised session after the end of their immobilization period (six weeks postoperatively). Additionally, the patients were instructed to perform the exercises once per week at home or at a gym on the weekends (never training on two consecutive days).

### 5. Adherence

At each group-based BFR-LLST added to usual care exercise supervised session two days a week (from week 7 – 12), the physical therapist registered, which exercises and how many repetitions, sets and the load the individual patient performed. Furthermore, the patients were asked to registerer number of sets, repetitions, and load of each exercise, when they performed the exercises at home, and bring the data to the following group session, so it could be registered by the physical terapist

Patients registered the date, cuff/elastic band, load, sets and number of repetitions and any complaints during the BFR-LLST at home in a training diary in the BFR-LLST leaflet (S4). A physical therapist registered the same data during the BFR-LLST supervised session at the SOS-R.

### 6. Motivation

Generally, the physical therapists encouraged patients to self-manage their rehabilitation and the importance of adhering to the BFR-LLST added to usual care exercise program, and through group dynamics from week 7 to week 12 postoperatively.

### 7. Progression decision rules

Exercises were progressed if patients were able to perform the number of repetitions maximum (RM) and form/quality described in the usual care exercise after cartilage and meniscus repair in the knee joint - week 7- postoperatively template (S6). If the patients were able to complete all three sets with correct form and quality, the patients were progressed to the next exercise level for the specific muscle group. The patients did not progress, if the number of repetitions were not reached or if form/quality were not obtained as described.

If patients were not able to perform the next exercise as described in the template, because of pain or lack of form, more resistance was added to the previous exercise, to ensure progression. The resistance applied in each exercise was registered and used in the next training session at home.

### 8. Exercise description

All patients had four different types of exercises during their scheduled nine weeks of rehabilitation period at SOS-R (Figure 1); 1) BFR-LLST supervised, which consisted of three individual and 12 group-based supervised sessions; 2) BFR-LLST home consisted of 30 home-based sessions; 3) Usual care exercise supervised was three individual and 12 group-based supervised sessions; and 4) Usual care exercise home consisted of 24 home-based sessions. After the BFR-LLST added to usual care exercise supervised intervention period, the physical therapists responsible for the usual care exercise supervised decided whether the patients should continue their rehabilitation.

The usual care exercise programs can be found in S5 and S6. The BFR-LLST protocol can be found in the table (see Table 1) and instructions are found in S4.

All exercises were illustrated with photographs and a thorough written description including number of sets, repetitions, and time under tension.

**Figure 1.**
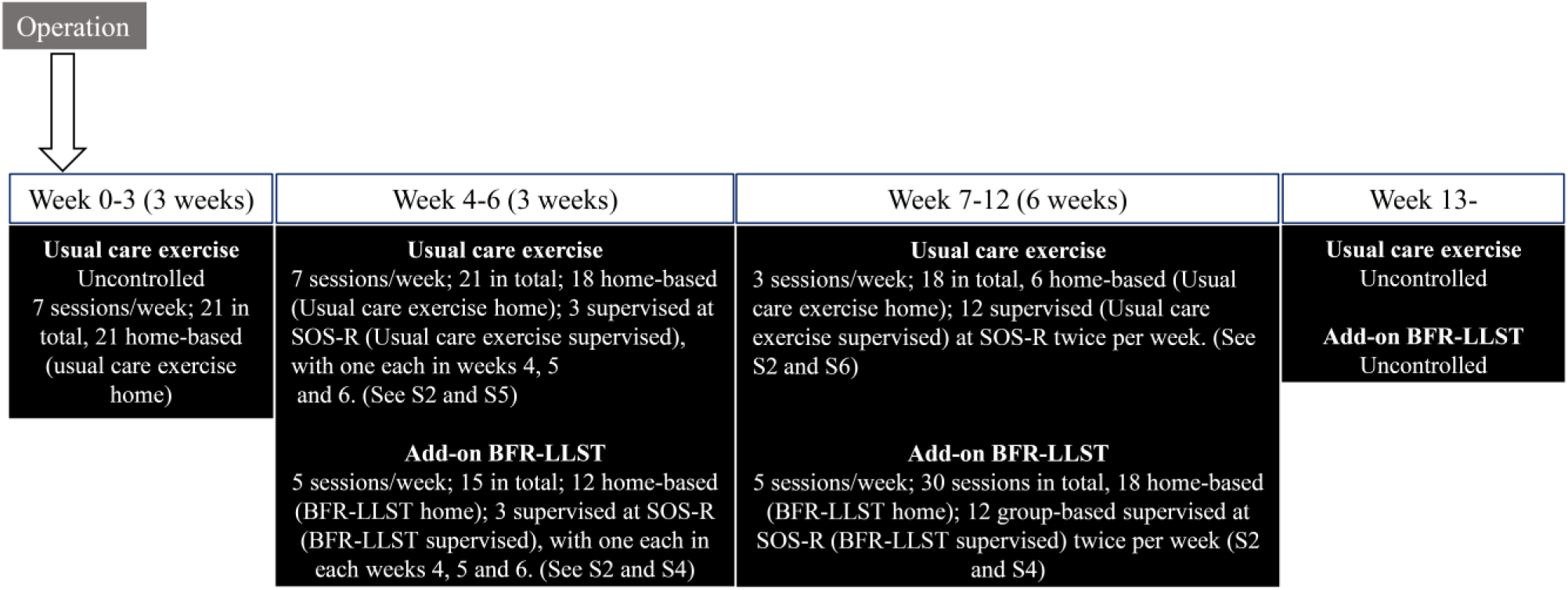

**Table 1.**
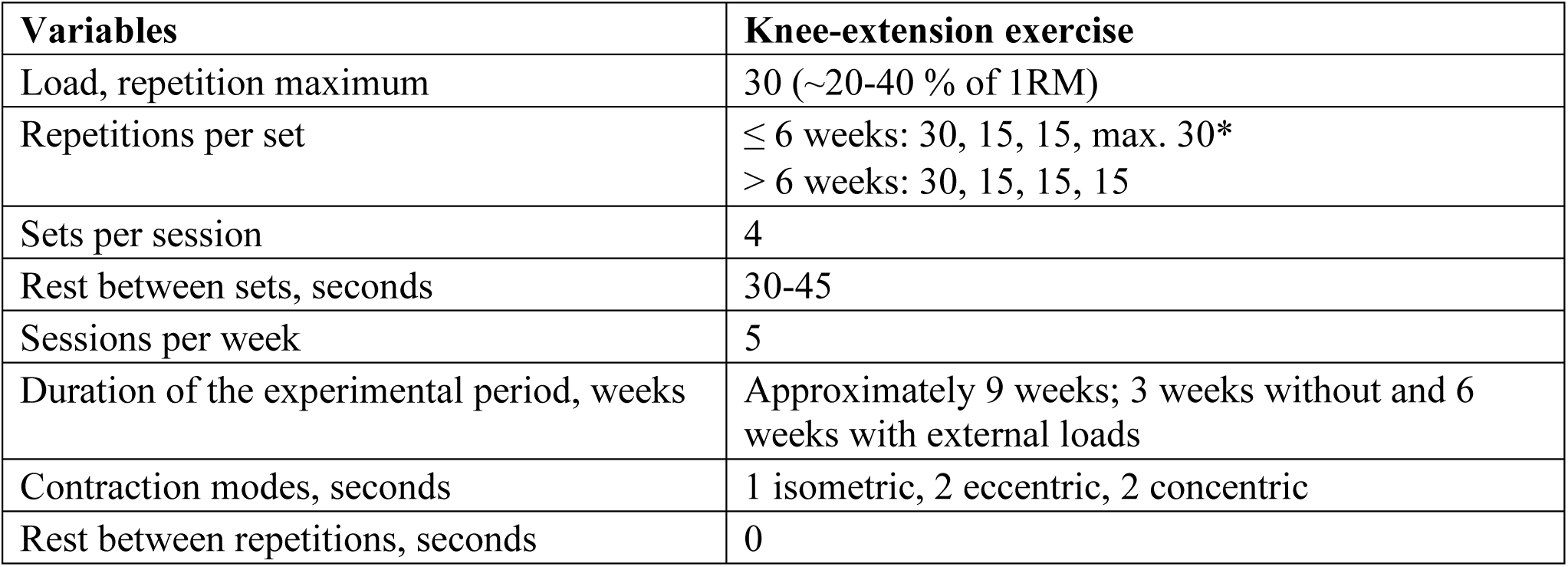
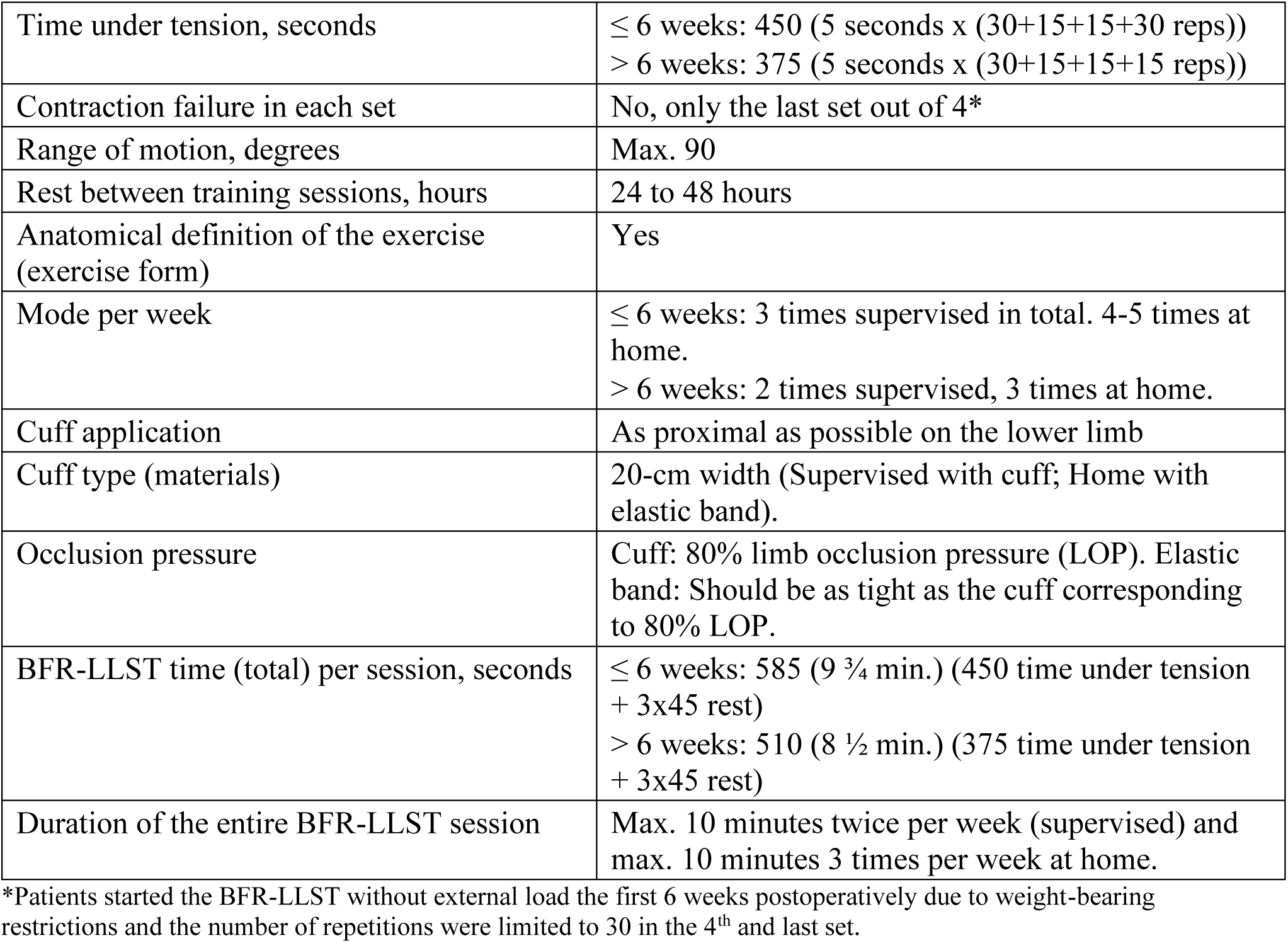
The BFR-LLST protocol for the knee-extension exercise

| Variables | Knee-extension exercise |
| --- | --- |
| Load, repetition maximum | 30 (~20-40 % of 1RM) |
| Repetitions per set | ≤ 6 weeks: 30, 15, 15, max. 30*<br>> 6 weeks: 30, 15, 15, 15 |
| Sets per session | 4 |
| Rest between sets, seconds | 30-45 |
| Sessions per week | 5 |
| Duration of the experimental period, weeks | Approximately 9 weeks; 3 weeks without and 6 weeks with external loads |
| Contraction modes, seconds | 1 isometric, 2 eccentric, 2 concentric |
| Rest between repetitions, seconds | 0 |
| S2 CERT (Consensus on Exercise Reporting Template) |  |
| Time under tension, seconds | ≤ 6 weeks: 450 (5 seconds x (30+15+15+30 reps))<br>> 6 weeks: 375 (5 seconds x (30+15+15+15 reps)) |
| Contraction failure in each set | No, only the last set out of 4* |
| Range of motion, degrees | Max. 90 |
| Rest between training sessions, hours | 24 to 48 hours |
| Anatomical definition of the exercise (exercise form) | Yes |
| Mode per week | ≤ 6 weeks: 3 times supervised in total. 4-5 times at home.<br>> 6 weeks: 2 times supervised, 3 times at home. |
| Cuff application | As proximal as possible on the lower limb |
| Cuff type (materials) | 20-cm width (Supervised with cuff; Home with elastic band). |
| Occlusion pressure | Cuff: 80% limb occlusion pressure (LOP). Elastic band: Should be as tight as the cuff corresponding to 80% LOP. |
| BFR-LLST time (total) per session, seconds | ≤ 6 weeks: 585 (9 ¾ min.) (450 time under tension + 3x45 rest)<br>> 6 weeks: 510 (8 ½ min.) (375 time under tension + 3x45 rest) |
| Duration of the entire BFR-LLST session | Max. 10 minutes twice per week (supervised) and max. 10 minutes 3 times per week at home. |

\*Patients started the BFR-LLST without external load the first 6 weeks postoperatively due to weight-bearing restrictions and the number of repetitions were limited to 30 in the 4<sup>th</sup> and last set.

### 9. Home program

Initially, all patients received a written description of the BFR-LLST leaflet (S4) explaining the advantages, disadvantages and potential risks related to the BFR intervention. Furthermore, the patients received hand-outs with two different home exercise programs for i) individual session week 3-6 postoperatively (S5), and ii) group session period week 7-12 postoperatively (S6), respectively. The exercise templates used during group-based usual care exercise supervised sessions were also used for the home training in the same period of as well as the following period from week 13.

### 10. Non-exercise components

At the first individual BFR-LLST added to usual care exercise session (baseline), the physical therapist instructed patients in correct gait patterns and how they should manage and control their knee joint pain and swelling. Other non-exercise components were non-standardized.

### 11. Adverse events

The BFR-LLST leaflet (S4) contained in-depth information about advantages, disadvantages and potential risks related to the BFR-LLST intervention. Additionally, any adverse event related to BFR-LLST at the supervised sessions or at home were reported by the patients to the physical therapist responsible for each BFR-LLST supervised session. If patients experienced clinical signs and symptoms of deep venous thrombosis at home, they were urged to contact a medical doctor or the physical therapist immediately (S4).

### 12. Setting

The exercises were performed at the supervised group-based classes twice a week at the Section for Orthopaedic and Sports Rehabilitation (SOS-R) Copenhagen, in the training hall. Home training were either performed in the patients’ own home or in a fitness center.

### 13.-15. Intervention description, Tailoring, Starting level

The usual care exercise after cartilage and meniscus repair in the knee joint - week 7postoperatively – program contained 44 exercises (S6). The selection of exercises for each patient was based on their strength and quality of exercise performance.

Exercises were progressed if patients were able to perform the number of repetitions maximum (RM) and form/quality described in the usual care (exercise) template, starting from the lowest level. If the patients were able to complete all three sets with correct form and quality, the patients were progressed to the next exercise level for the specific muscle group. The patients did not progress, if the number of repetitions were not reached or if form/quality were not obtained as described.

If patients were not able to perform the next exercise as described in the template, because of pain or lack of strength, more resistance was added to the previous exercise, to ensure progression. The resistance applied in each exercise was registered and was used in the next training session at home.

Each BFR-LLST added to usual care exercise group-based session was initiated by ten minutes of pre-warming up exercises. Then 30 to 35 minutes were spent on exercises from the template, and the remaining time (10 to 15 minutes) was spent on BFR-LLST according to the protocol.

After the six weeks (week 7-12) with group-based usual care exercise supervised, the patients were instructed to perform the exercises three times per week. During the supervised rehabilitation phase, they were taught how to progress the exercises from the usual care exercise program.

The patients could experience acceptable pain during exercise. The pain needed to resolve immediately after performing the exercise (within 24 hours). The patients were instructed in differentiating between ‘good’ pain (muscle soreness/pain or pain around the knee) and ‘bad’ pain (pain inside the knee).

### 16. Fidelity to the intervention plan

At each BFR-LLST added to the usual care exercise session, number of training sessions performed for BFR-LLST (supervised and home), usual care exercise (supervised and at home), the BFR-LLST descriptors of limb occlusion pressure applied, number of sets, repetitions and external load lifted were recorded. BFR-LLST descriptors at home were patient-reported via a training diary (S4). Furthermore, the specific exercises performed at each usual care exercise supervised session were noted.

Adherence to the programs was acceptable with patients performing, on average, more than 80% of the scheduled BFR-LLST added to the usual care exercise sessions both supervised and at home were performed (S7 Table 2).

## S3 Rehabilitation regimens - Cartilage (Steadman procedure) and meniscus repair

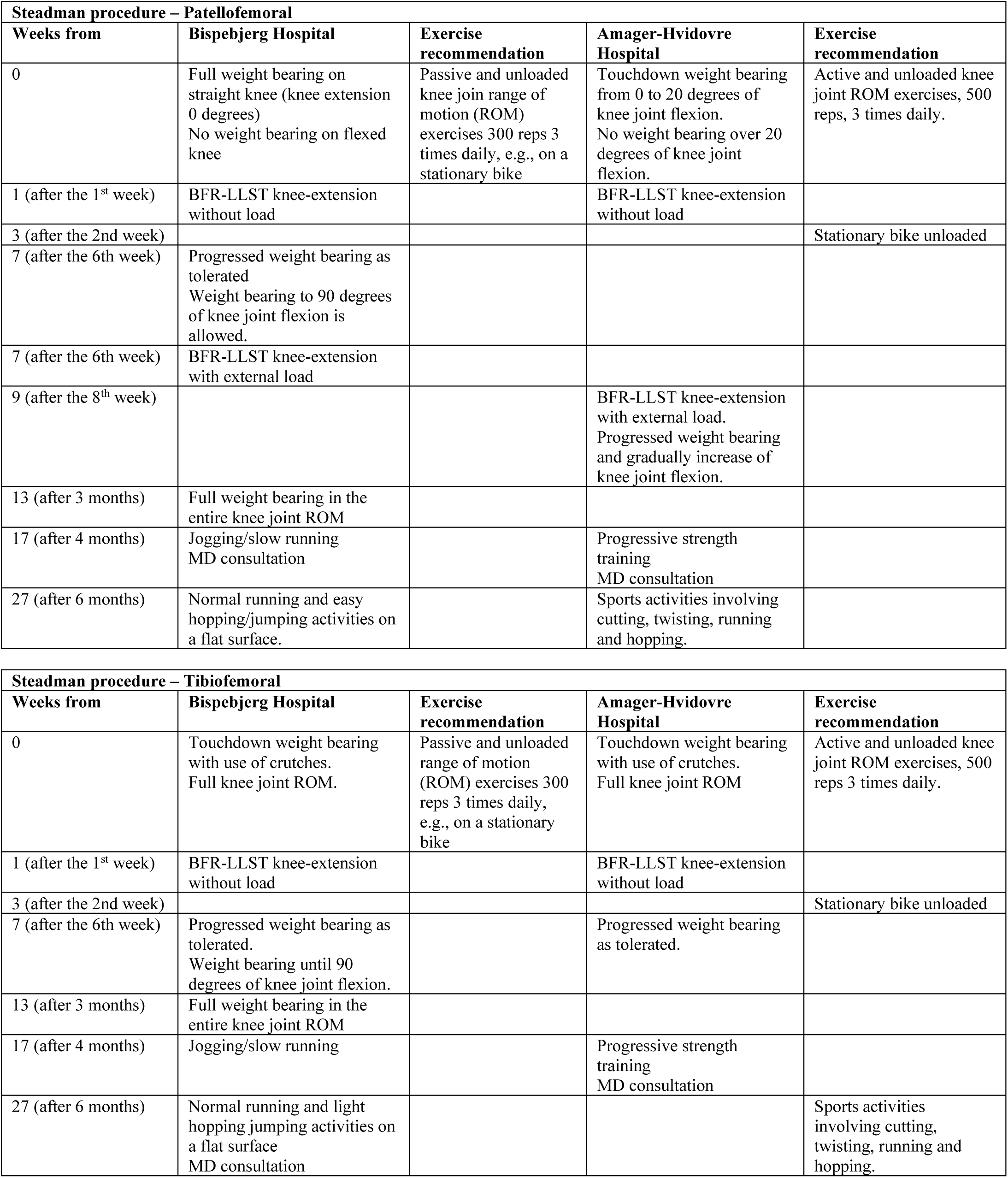
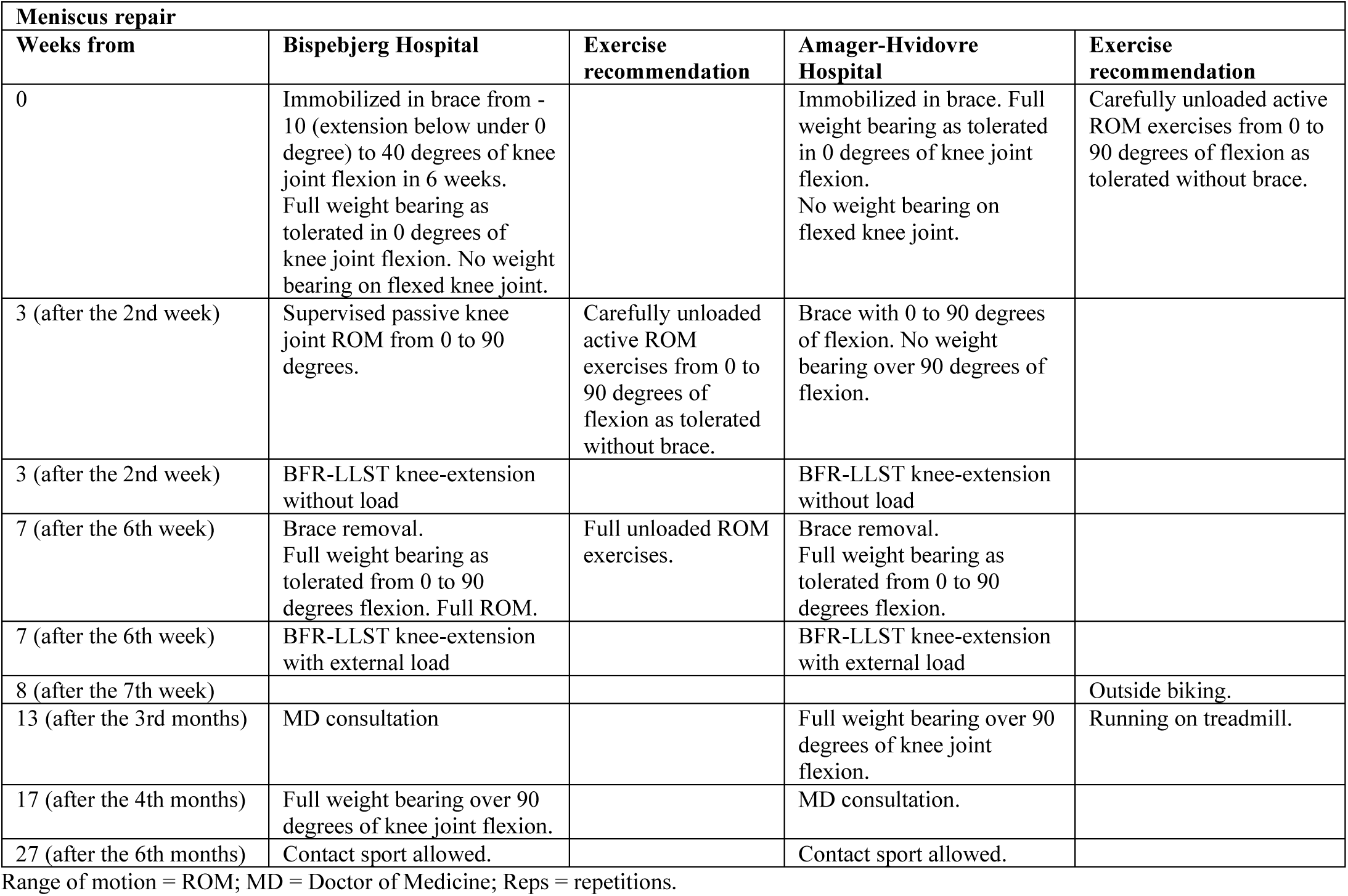

## Blood flow restriction exercise

*Example of Blood flow restriction exercise. medRxiv policy does not allow for publication of images that may be identifi-able to the individual, their friends, family, or neighbors. Pictures are available from the corresponding author and from the peer-reviewed paper, once published.*

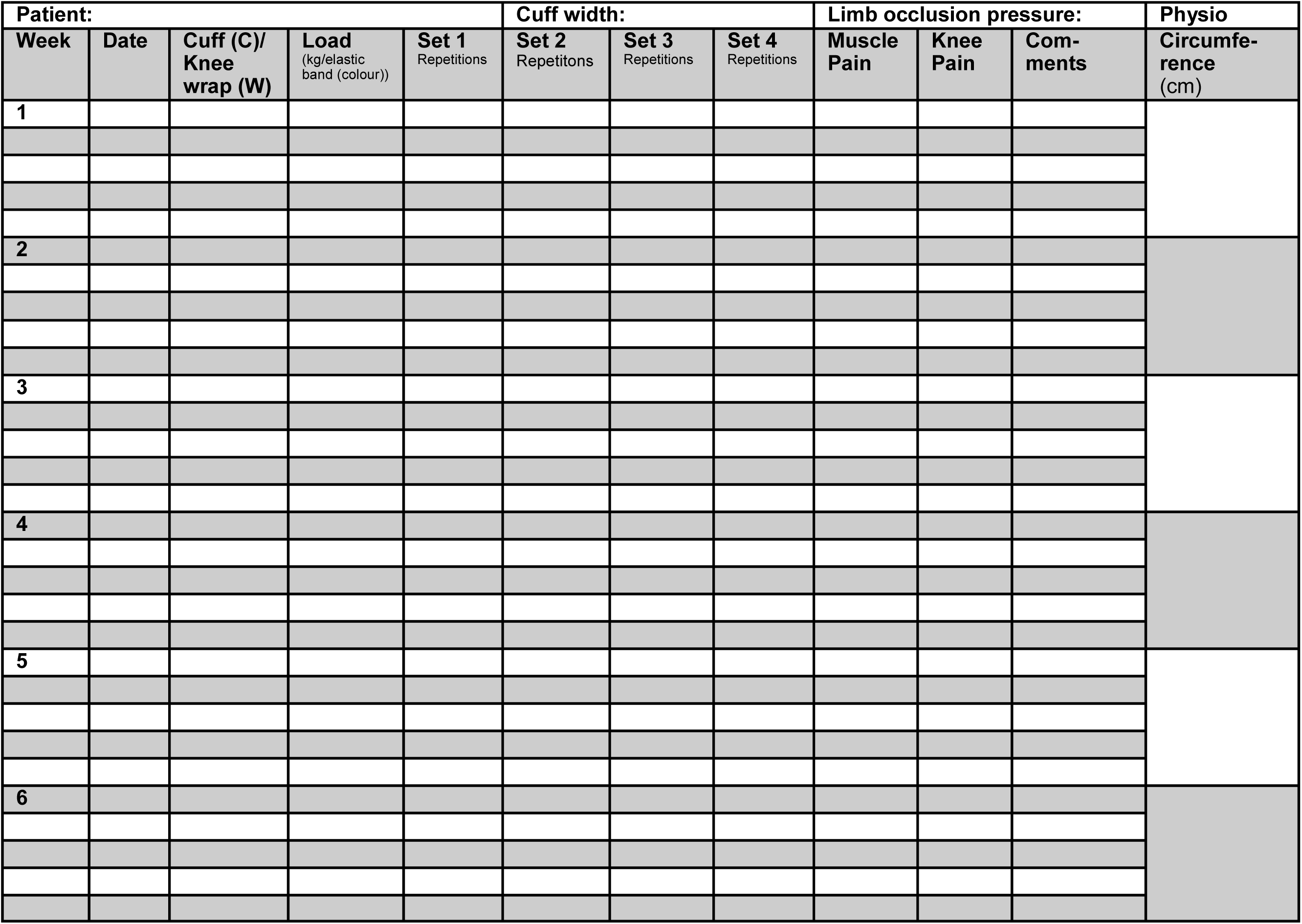

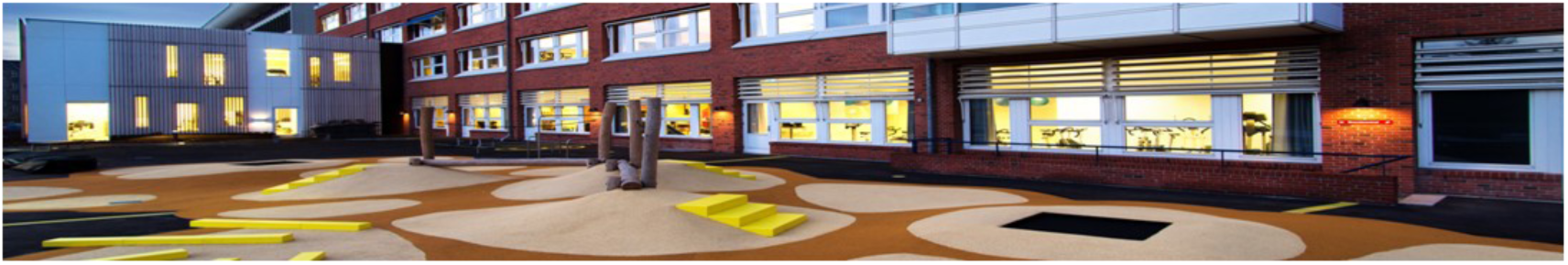

### Blood flow restriction exercise (BFR)

Blow flow restriction exercise (BFR) is a form of training in which the blood flow is restricted by an inflatable cuff or elastic band at the top of the thigh. This type of exercise has been found to increase muscle strength approximately to the same degree as traditional strength training. The advantage is that you can exercise intensively without high loads. When you are allowed to fully load your leg and experience no or mild knee pain, we recommend that you start traditional strength training.

### Can I perform BFR exercise?

□ Standardized BFR exercise is safe. As we limit blood flow to the leg, we do not recommend BFR exercise if you:
□ Have high blood pressure (hypertension), diabetes, and/or get bruising easily.
□ Have problems with your heart or blood vessels (such as atherosclerosis).
□ Have a neurological disease.
□ Have a psychiatric diagnosis that prevents you from exercising regularly.
□ Have an abuse of alcohol, drugs or euphoric drugs.
□ Have cancer
□ Have an active inflammatory state in the body (septic active infection)
□ Is pregnant

### BFR exercise in practice

The inflatable cuff is placed as far up on your thigh as possible.

*Example of placement of the inflatable cuff. medRxiv policy does not allow for publication of images that may be identifiable to the individual, their friends, family, or neighbors. Pictures are available from the corresponding author and from the peer-reviewed paper, once published*.

The physical therapist inflates the cuff to find the right pressure. Most often, the pressure will be between 80 and 140 mmHg (millimeters of mercury).

If you are using an elastic band, put the entire band around your thigh. It is placed in the same place as the cuff. You should experience the same pressure in your thigh as when using the cuff.

*Example of placement of the the elastic band. medRxiv policy does not allow for publication of images that may be identifiable to the individual, their friends, family, or neighbors. Pictures are available from the corresponding author and from the peer-reviewed paper, once published*.

If you are using an elastic belt, put the entire belt around your thigh. It is placed in the same place as the cuff. You should experience the same pressure in your thigh as when using the cuff.

You have to complete 4 sets..

1. set: 30 repetitions, ½ minute break
2. set: 15 repetitions, ½ minute break
3. set: 15 repetitions, ½ minute break
4. set: 15 repetitions (Maximum 30 repetitions)

When performing a repetition, it should take 2 seconds to extend the leg to a horizontal position, 1 second to hold the leg fully horizontal, 2 seconds to lower the leg again, and 1 second to relax with the leg in a vertical position.

*Do not restrict the thigh for more than 10 minutes while exercising*.

### Load when performing BFR exercise

Start training without load. In the 4^th^ and last set, perform as many repetitions as you can - however, no more than 30 repetitions. When you are allowed to load your knee, it is important that you gradually increase your exercise load. When you manage more than 2 repetitions on top of the 15 repetitions in your 4th and last set, the load should be increased in the next workout (+2 principle).

We recommend that you BFR exercise 5 times a week. It is an advantage to finish with the BFR exercise, when you train.

On the last page of the leaflet, there is a training diary, you are obliged to fill out. Then you and your physical therapist can follow your training. In this way we make sure that you train optimally.

If you want to train your posterior thigh muscle, the same principles apply as when you are training the anterior thigh muscle. The starting position is of course different. If you want to exercise both the anterior and posterior thigh muscles, there must be a 5-minute break in between your workouts.

### Good advice

The physical therapist determines the pressure you need to work out with. The cuff or band you are training with should not squeeze more than a 7 on a scale of 0 (no pressure in the muscle) to 10 (unbearable pressure in the muscle).

When you exercise, you will experience pain/discomfort in your muscle, but you must avoid pain in the knee joint. On a scale of 0 (no pain) to 10 (unbearable pain), avoid experiencing pain more than 3 out of 10 in the knee joint.

You will sometimes notice that your leg becomes discoloured when performing BFR exercise. These symptoms are natural, and will disappear when you do not squeeze (restrict) your leg any more. I

If you find that the leg is red all the time, it hurts, is swollen and you have a fever, you should contact a doctor or the treating physical therapist immediately.

Once you have completed your BFR exercise, loosen the cuff/elastic band little by little. You may feel dizzy when you loosen your band. If you feel very dizzy, lie down with your legs elevated.

Elastic wide bands (knee wraps) and inflatable cuffs are available in specialized stores. We recommend that you contact your physical therapist before purchasing.

## S5 REHABILITATION PROGRAM Cartilage repair procedure knee (tibiofemoral) (After 14 days and until week 6 incl.)

The exercise program is performed daily

*Example of patient doing active knee extension. medRxiv policy does not allow for publication of images that may be identifiable to the individual, their friends, family, or neighbors. Pictures are available from the corresponding author and from the peer-reviewed paper, once published.*

**<u>1. Active knee extension</u>**

Sit with extended leg on the floor or exercise mat. Place a firm cushion under the knee.

Tighten up the front thigh muscle, so the heel is lifted from the floor to full knee extension. Hold the tension for 5 sec. then slowly lower the heel again.

**3 sets of 15 repetitions**

*Example of patient doing unloaded knee flexion. medRxiv policy does not allow for publication of images that may be identifiable to the individual, their friends, family, or neighbors. Pictures are available from the corresponding author and from the peer-reviewed paper, once published.*

**<u>2. Unloaded knee flexion</u>**

Sit with the leg resting on a smooth surface and the foot placed on a piece of cloth. Then bend the knee, put your hands around the leg and pull your foot as far up towards the back of your thigh, while the foot slides on the surface. Then let the foot slide back in a controlled manner to the extended starting position.

**5 sets of 60 repetitions**

If you can flex more than 95 degrees in the knee, you replace the exercise with the exercise bike without resistance for 10 min.

*Example of patient doing passive knee extension. medRxiv policy does not allow for publication of images that may be identifiable to the individual, their friends, family, or neighbors. Pictures are available from the corresponding author and from the peer-reviewed paper, once published.*

**<u>3. Passive knee extension</u>**

Sit with fully extended knee on the floor. Extend the knee by the aid of a solid rope/strap, so that the heel is lifted from the surface and the knee is fully extended. Hold the extension for 5 sec., and then slowly lower the heel.

**3 sets of 15 repetitions**

*Example of patient doing prone knee flexion. medRxiv policy does not allow for publication of images that may be identifiable to the individual, their friends, family, or neighbors. Pictures are available from the corresponding author and from the peer-reviewed paper, once published.*

**<u>4. Prone knee flexion</u>**

Lie in a prone position with fully extended knees. Bend the knee, so the heel is moved towards the buttock. Avoid swaying in the lower back, when the heel is pulled towards the buttock.

To increase the difficulty, put an elastic band around the ankles as resistance.

**3 sets of 15 repetitions**

*Example of patient doing side laying stretched leg lift. medRxiv policy does not allow for publication of images that may be identifiable to the individual, their friends, family, or neighbors. Pictures are available from the corresponding author and from the peer-reviewed paper, once published.*

**<u>5. Side laying stretched leg lift</u>**

Lie on your side with the lower leg slightly bend to maintain optimal balance.

Lift the upper leg upwards and slanting backwards.

Slowly lower the leg again. It is important that you maintain a straight line between hip, knee and ankle during the entire movement.

**3 sets of 15 repetitions**

## S5 REHABILITATION PROGRAM Cartilage repair procedure knee (patellofemoral) (After 14 days and until week 6 incl.)

The exercise program is performed daily

*Example of patient doing active knee extension. medRxiv policy does not allow for publication of images that may be identifiable to the individual, their friends, family, or neighbors. Pictures are available from the corresponding author and from the peer-reviewed paper, once published*.

**<u>1. Active knee extension</u>**

Sit with extended leg on the floor or mat. Place a firm cushion in the popliteal. Tighten up the front thigh muscle, so the heel is lifted from the floor to full knee extension.

Hold the tension for 5 sec. then slowly lower the heel again.

**3 sets of 15 repetitions**

*Example of patient doing unloaded knee flexion. medRxiv policy does not allow for publication of images that may be identifiable to the individual, their friends, family, or neighbors. Pictures are available from the corresponding author and from the peer-reviewed paper, once published*.

**<u>2. Unloaded knee flexion</u>**

Sit with the leg resting on a smooth surface and the foot placed on a piece of cloth. Then bend the knee, using the hands while the foot slides on the surface, then let the foot slide controlled back to extended starting position.

**5 sets of 60 repetitions**

*Example of patient doing passive knee extension. medRxiv policy does not allow for publication of images that may be identifiable to the individual, their friends, family, or neighbors. Pictures are available from the corresponding author and from the peer-reviewed paper, once published*.

**<u>3. Passive knee extension</u>**

Sit with extended leg on the floor or other firm surface.

Extend the knee by the aid of a solid rope, so that the heel is lifted from the surface and the knee is fully extended. Hold the extension for 5 secs., and then slowly lower the heel.

**3 sets of 15 repetitions**

*Example of patient doing prone knee flexion. medRxiv policy does not allow for publication of images that may be identifiable to the individual, their friends, family, or neighbors. Pictures are available from the corresponding author and from the peer-reviewed paper, once published*.

**<u>4. Prone knee flexion</u>**

Lie in a prone position with fully extended knees. Bend one knee, so the heel is moved towards the buttock. Avoid swaying in the lower back, when the heel is pulled towards the buttock.

To increase the difficulty, put an elastic band around the ankles as resistance.

**3 sets of 15 repetitions**

*Example of patient doing side laying stretched leg lift. medRxiv policy does not allow for publication of images that may be identifiable to the individual, their friends, family, or neighbors. Pictures are available from the corresponding author and from the peer-reviewed paper, once published*.

**<u>5. Side laying stretched leg lift</u>**

Lie on the side, with the lower leg slightly bend to maintain optimal balance.

Lift the upper leg upwards and slanting backwards.

**3 sets of 15 repetitions**

*Example of patient doing standing heel lift. medRxiv policy does not allow for publication of images that may be identifiable to the individual, their friends, family, or neighbors. Pictures are available from the corresponding author and from the peer-reviewed paper, once published*.

**<u>6. Standing heel lift</u>**

Stand on your forefoot with extended knees on a step with the heels slightly hanging off. Raise your body, so you are standing on your forefoot. Hold the position for 5 sec.

Then slowly lower the heels.

**3 sets of 15 repetitions**

## S5 REHABILITATION PROGRAM Meniscus repair (After 14 days and until week 6 incl.)

The exercise program is performed daily

**<u>1. Active knee extension</u>**

Hold the tension for 5 sec. then slowly lower the heel again.

**3 sets of 15 repetitions**

**<u>2. Unloaded knee flexion</u>**

**Attention: Max. 90 degrees of flexion! 5 sets of 60 repetitions**

**<u>3. Passive knee extension</u>**

Sit with the leg on the floor or other firm surface. Extend the knee by the aid of a solid rope, so that the heel is lifted from the surface and the knee is fully extended. Hold the extension for 5 secs., and then slowly lower the heel.

**3 sets of 15 repetitions**

**<u>4. Prone knee flexion</u>**

To increase the difficulty, put an elastic band around the ankles as resistance.

**Attention: Max. 90 degrees of flexion! 3 sets of 15 repetitions**

**<u>5. Side laying stretched leg lift</u>**

Lie on your side with the lower leg slightly bend to maintain optimal balance.

Lift the upper leg upwards and slanting backwards.

**3 sets of 15 repetitions**

**<u>6. Standing heel lift</u>**

Stand with extended knees on the floor or a small step with the heels over the edge.

Raise the body, so you are standing on your toes. Hold the tension for 5 sec.

Then slowly lower the heels towards the floor.

**3 sets of 15 repetitions**

## S6 Usual care exercise after cartilage and meniscus repair in the knee joint - week 7 postoperative

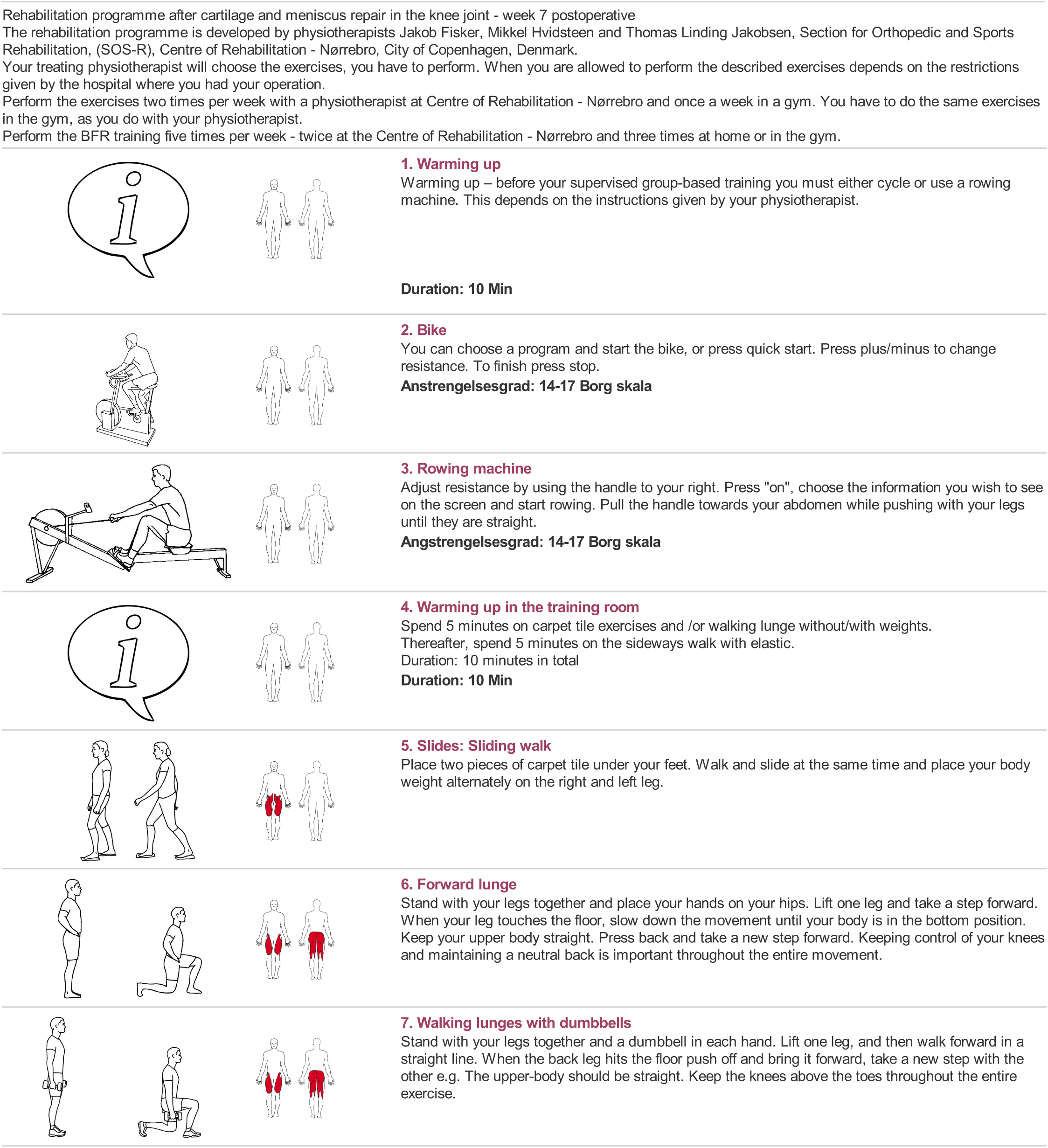
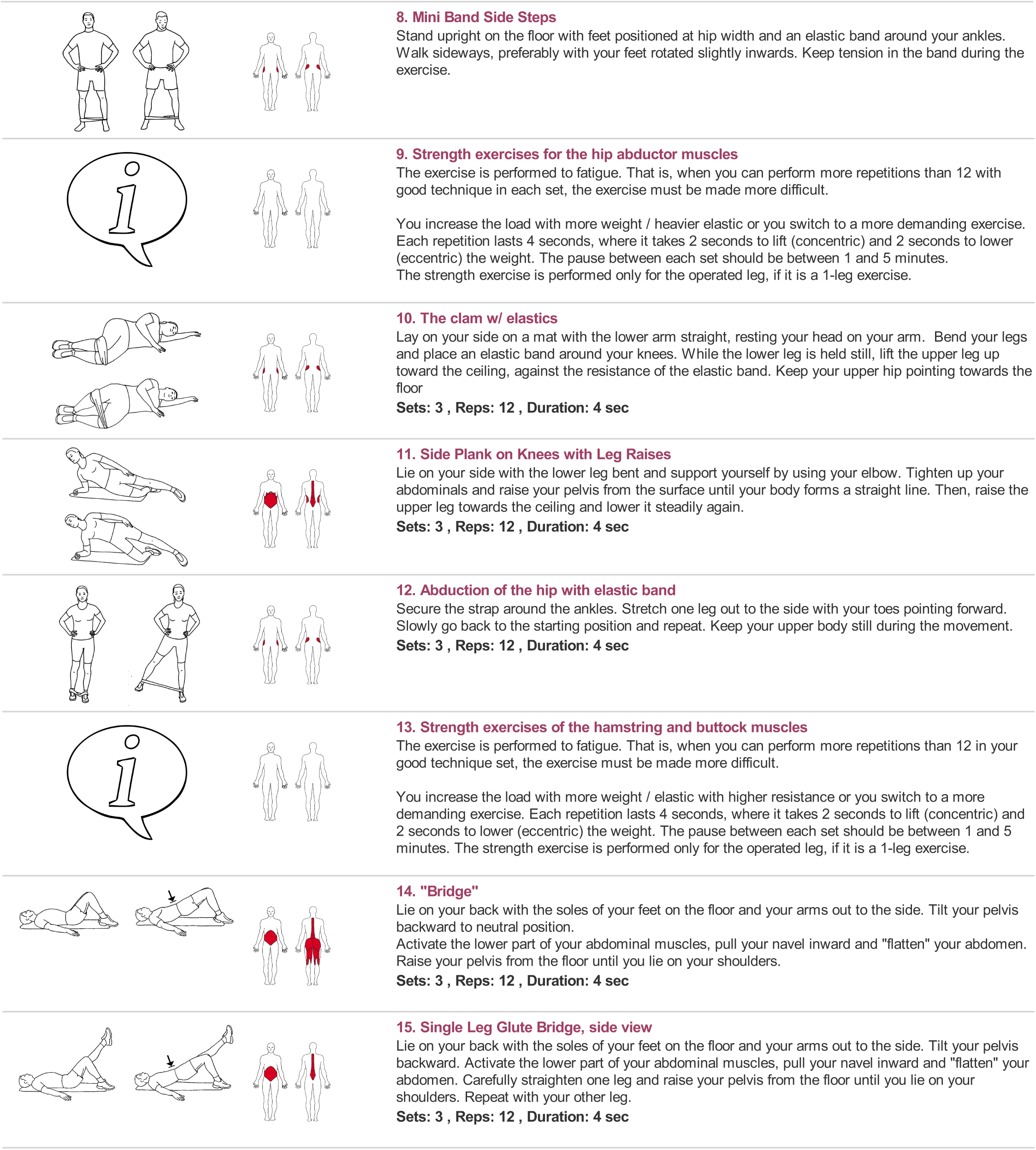
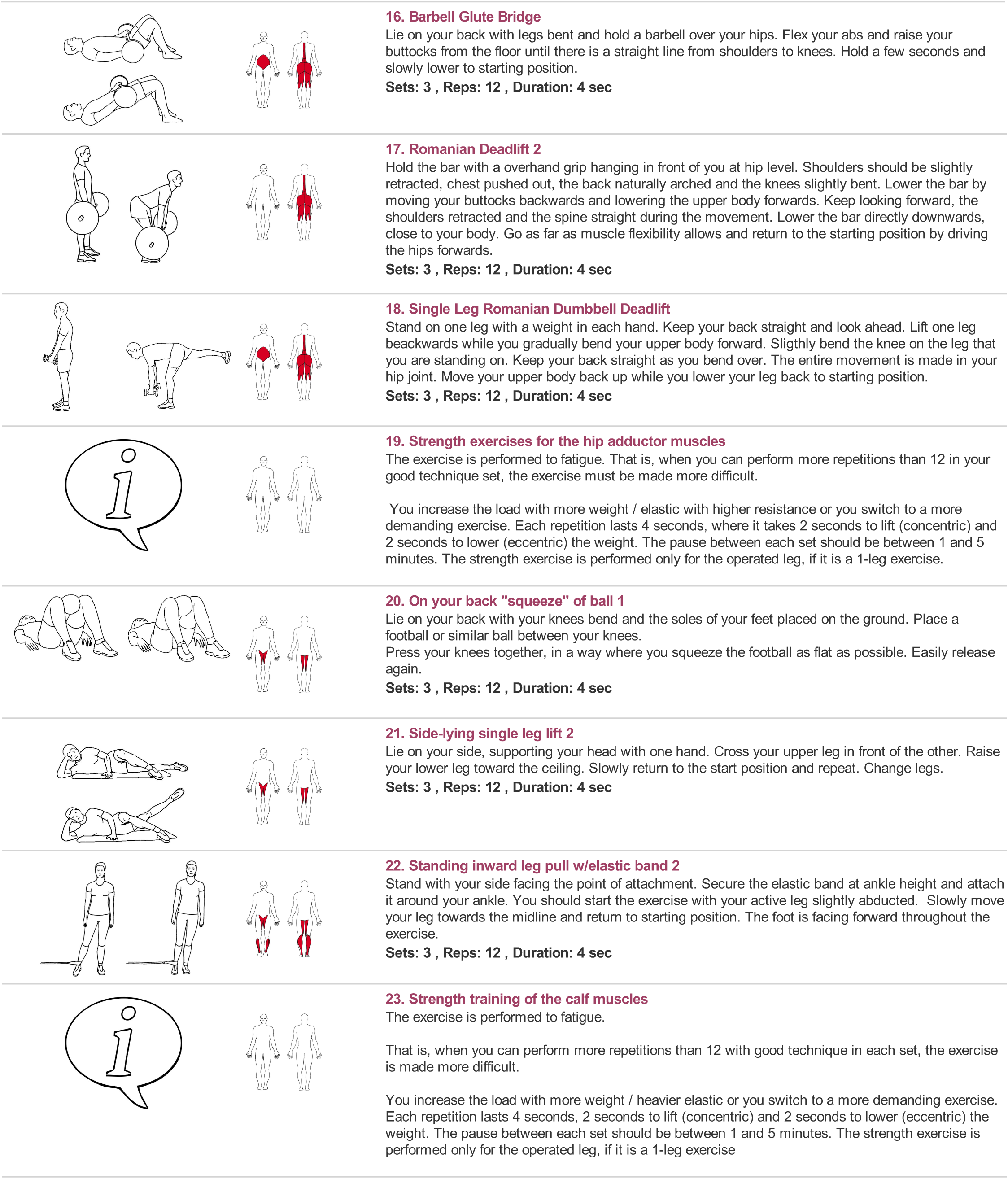
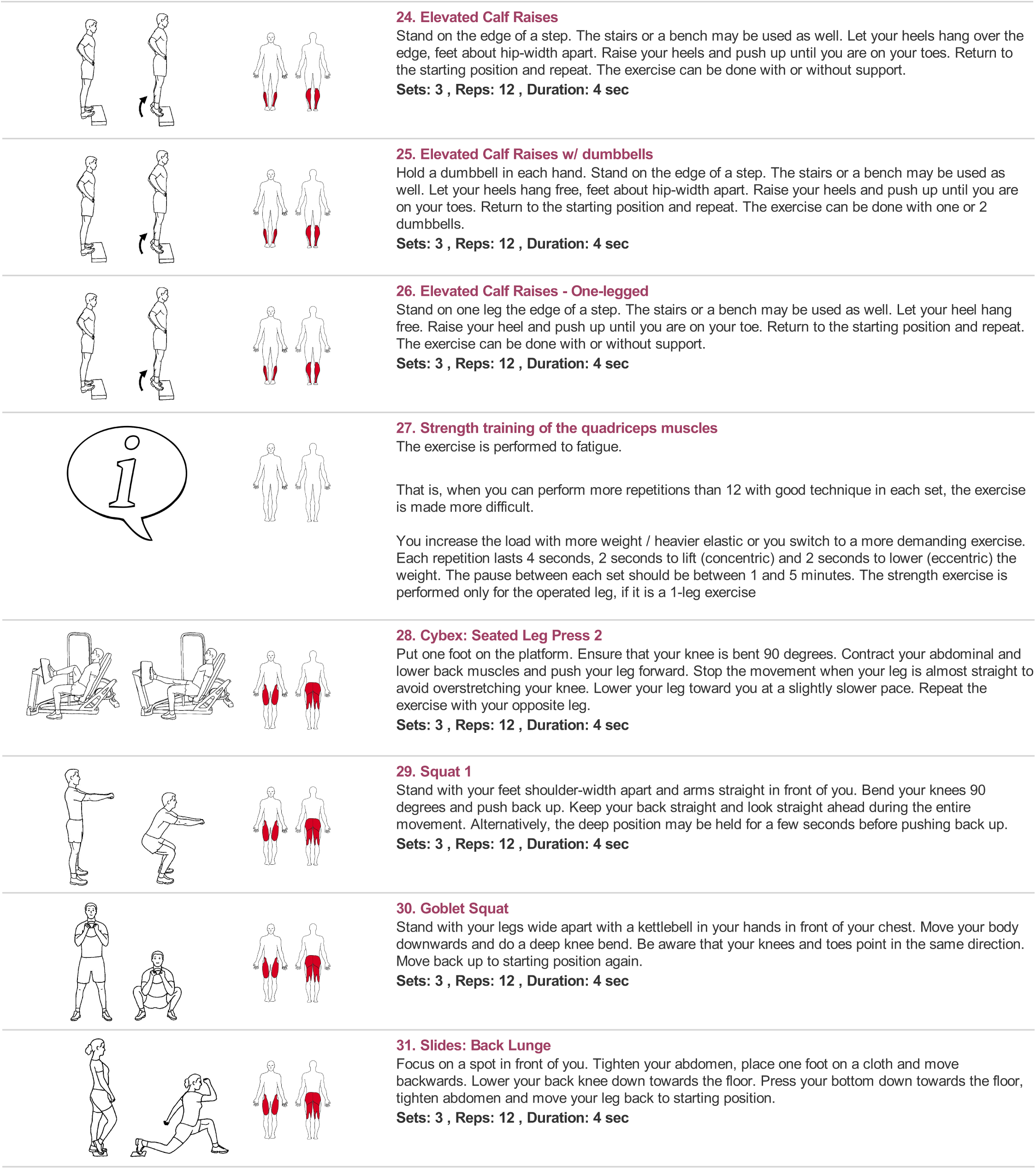
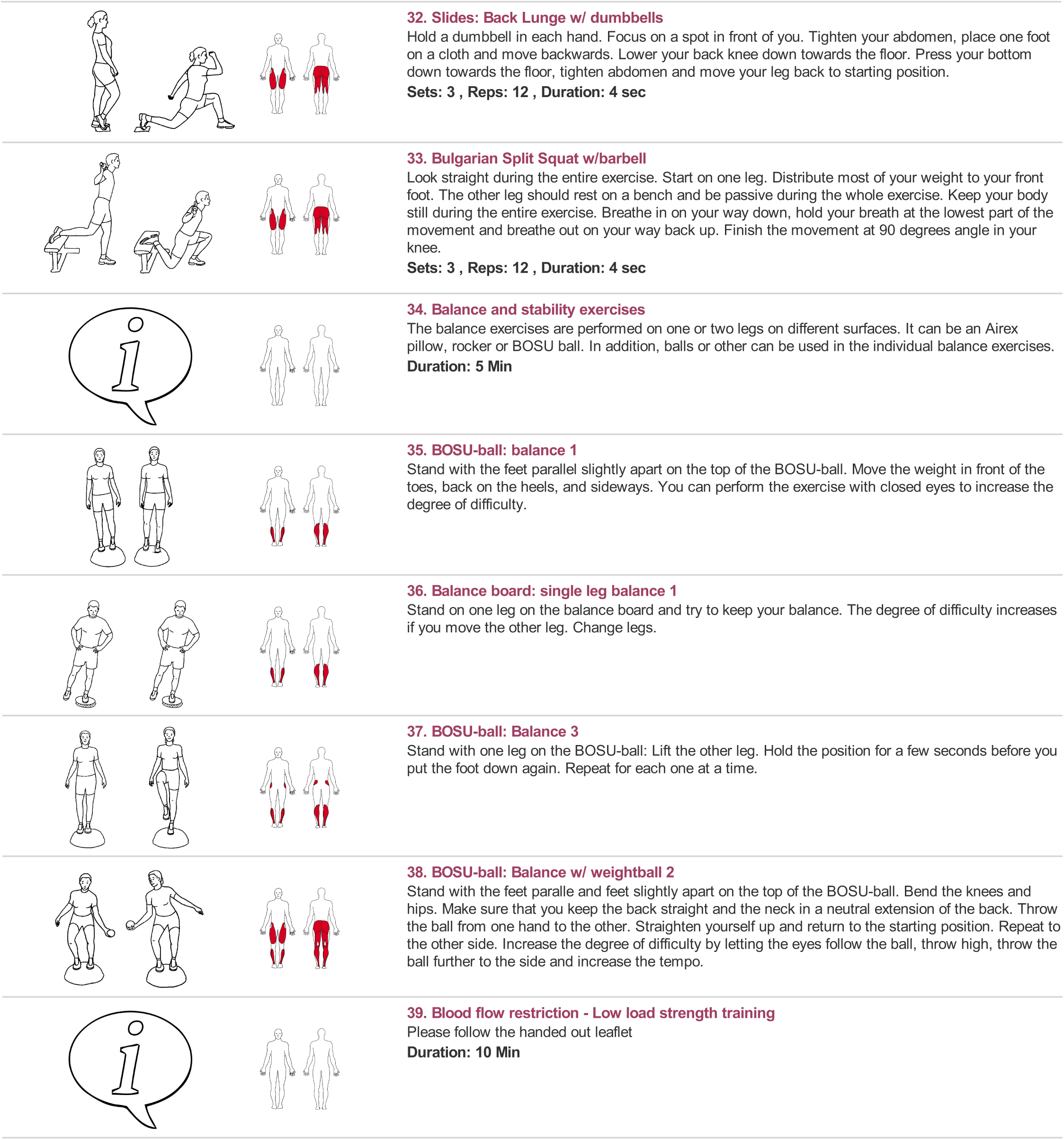
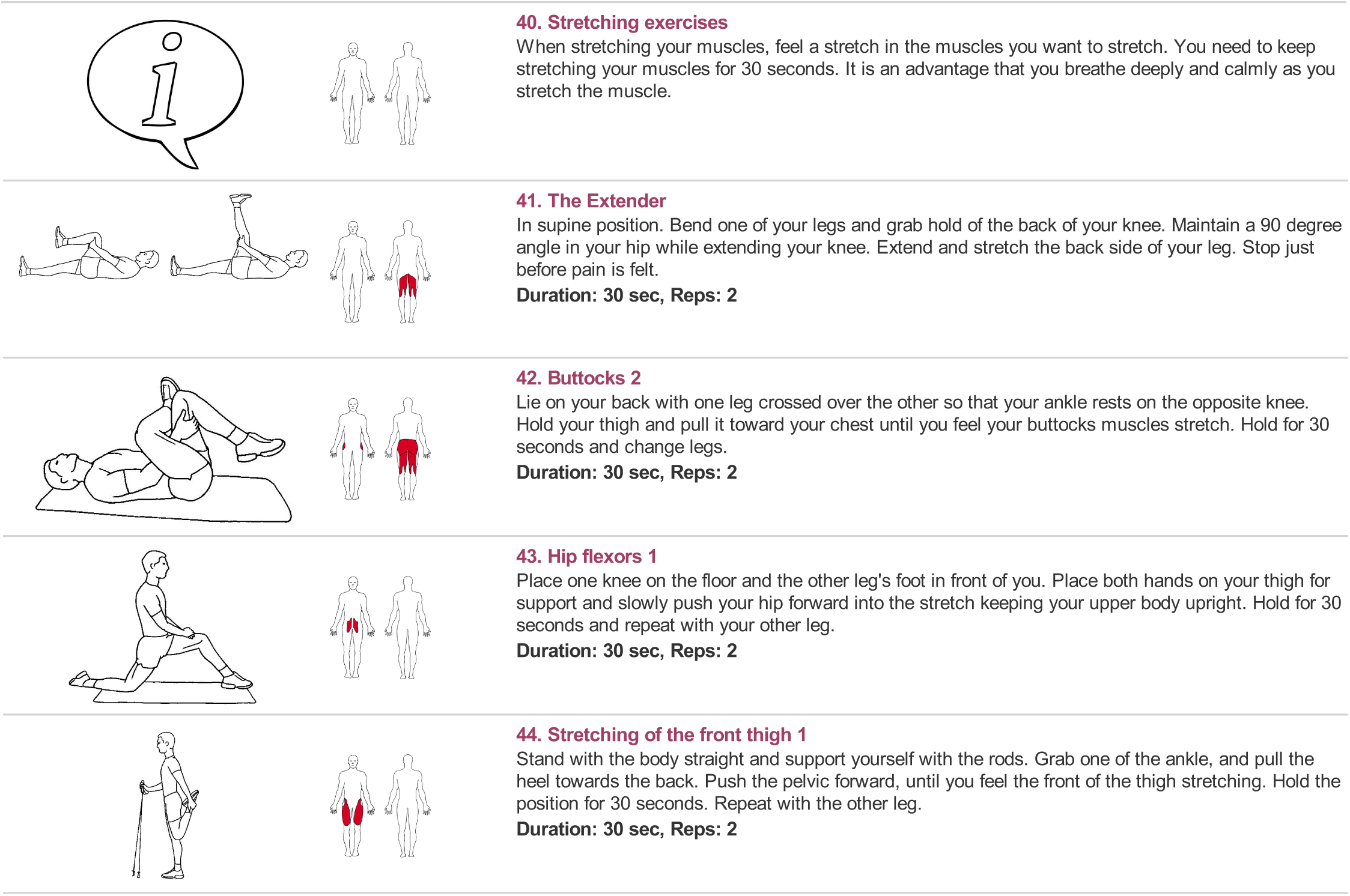

## S7 Supplementary Results Content

**S7 Table 1.**
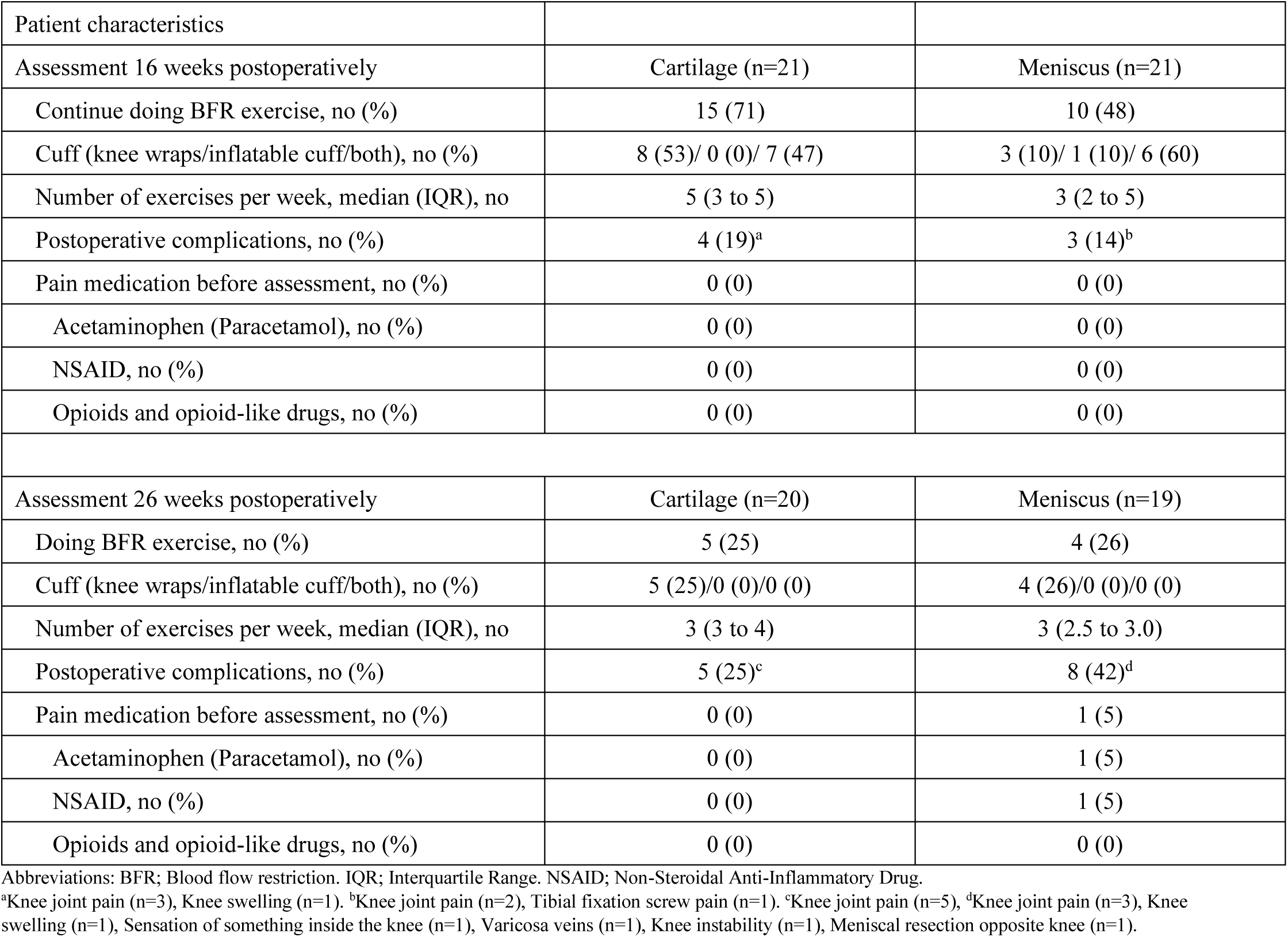
Patient characteristics at 16- and 26-week assessment

| Patient characteristics |  |  |
| --- | --- | --- |
| Assessment 16 weeks postoperatively | Cartilage (n=21) | Meniscus (n=21) |
| Continue doing BFR exercise, no (%) | 15 (71) | 10 (48) |
| Cuff (knee wraps/inflatable cuff/both), no (%) | 8 (53)/ 0 (0)/ 7 (47) | 3 (10)/ 1 (10)/ 6 (60) |
| Number of exercises per week, median (IQR), no | 5 (3 to 5) | 3 (2 to 5) |
| Postoperative complications, no (%) | 4 (19)<sup>a</sup> | 3 (14)<sup>b</sup> |
| Pain medication before assessment, no (%) | 0 (0) | 0 (0) |
| Acetaminophen (Paracetamol), no (%) | 0 (0) | 0 (0) |
| NSAID, no (%) | 0 (0) | 0 (0) |
| Opioids and opioid-like drugs, no (%) | 0 (0) | 0 (0) |
| Assessment 26 weeks postoperatively | Cartilage (n=20) | Meniscus (n=19) |
| Doing BFR exercise, no (%) | 5 (25) | 4 (26) |
| Cuff (knee wraps/inflatable cuff/both), no (%) | 5 (25)/0 (0)/0 (0) | 4 (26)/0 (0)/0 (0) |
| Number of exercises per week, median (IQR), no | 3 (3 to 4) | 3 (2.5 to 3.0) |
| Postoperative complications, no (%) | 5 (25)<sup>c</sup> | 8 (42)<sup>d</sup> |
| Pain medication before assessment, no (%) | 0 (0) | 1 (5) |
| Acetaminophen (Paracetamol), no (%) | 0 (0) | 1 (5) |
| NSAID, no (%) | 0 (0) | 1 (5) |
| Opioids and opioid-like drugs, no (%) | 0 (0) | 0 (0) |

Abbreviations: BFR; Blood flow restriction. IQR; Interquartile Range. NSAID; Non-Steroidal Anti-Inflammatory Drug.
<sup>a</sup>Knee joint pain (n=3), Knee swelling (n=1). <sup>b</sup>Knee joint pain (n=2), Tibial fixation screw pain (n=1). <sup>c</sup>Knee joint pain (n=5), <sup>d</sup>Knee joint pain (n=3), Knee swelling (n=1), Sensation of something inside the knee (n=1), Varicosa veins (n=1), Knee instability (n=1), Meniscal resection opposite knee (n=1).

**S7 Table 2.** Clinical application (adherence), training characteristics and pain at rest and during BFR-LLST added to usual care exercise

|  | Cartilage (n=21) | Meniscus (n=21) | Total (n=42) |
| --- | --- | --- | --- |
| **Clinical application (adherence)<sup>a</sup>** |  |  |  |
| BFR-LLST supervised, mean 1±SD (range, % completed), sessions | 13 ± 1 (9-15, 84) | 13 ± 2 (9-15, 84) | 13 ± 2 (9-15, 84) |
| BFR-LLST home, mean 1±SD (range, % completed), sessions | 37 ± 9 (21-57, 124) | 34 ± 5 (26-43, 113) | 35 ± 8 (21-57, 118) |
| BFR-LLST total, mean 1±SD (range, % completed), sessions | 50 ± 10 (34-70, 110) | 47 ± 6 (36-58, 107) | 48 ± 8 (31-70, 107) |
| Usual care exercise supervised, mean 1±SD (range, % completed), sessions | 13 ± 1 (9-15, 84) | 13 ± 2 (9-15, 84) | 13 ± 2 (9-15, 84) |
| Usual care exercise home, mean 1±SD (range, % completed), sessions | 31 ± 13 (14-61, 94) | 29 ± 12 (1-48, 97) | 30 ± 12 (1-61, 102) |
| Usual care exercise total, mean 1±SD (range, % completed), sessions | 43 ± 12 (26-74, 92) | 42 ± 12 (13-63, 94) | 42 ± 12 (13-74, 98) |
| **Training characteristics** |  |  |  |
| Limb occlusion pressure (LOP), mean ±1SD (range), mmHg<sup>b</sup> | 188 ± 17 (160-240) | 187 ± 22 (150-240) | 187 ± 19 (150-240) |
| Relative applied 80% limb occlusion pressure (LOP), mean ±1SD (range), mmHg<sup>c</sup> | 135 ± 14 (112-176) | 134 ± 17 (104-176) | 134 ± 15 (104-176) |
| Number of repetitions per BFR-LLST session, mean ±1SD (range), no | 82 ± 12 (15-136) | 81 ± 12 (10-135) | 81 ± 12 (10-136) |
| Performed the recommended number of repetitions per BFR-LLST session (min. 75 repetitions), mean ± 1SD, % | 80 ± 4 | 78 ± 4 | 79 ± 4 |
| External training load during BFR, median (IQR, range), kg | 1 (0 to 2, 0-7) | 1 (0 to 2, 0-5) | 1 (0 to 2, 0-7) |
| **Knee joint and quadriceps pain** |  |  |  |
| Knee joint pain at rest before BFR-LLST supervised session, median (IQR, range), VAS-mm | 0 (0 to 0, 0-61) | 0 (0 to 1, 0-70) | 0 (0 to 0, 0-70) |
| Knee joint pain at rest after BFR-LLST supervised session, median (IQR, range), VAS-mm | 0 (0 to 0, 0-65) | 0 (0 to 0, 0-49) | 0 (0 to 0, 0-65) |
| Quadriceps muscle pain at rest before BFR-LLST supervised session, median (IQR, range), VAS-mm | 0 (0 to 0, 0-51) | 0 (0 to 0, 0-40) | 0 (0 to 0, 0-51) |
| Quadriceps muscle pain at rest after BFR-LLST supervised session, median (IQR, range), VAS-mm | 0 (0 to 13, 0-70) | 0 (0 to 9, 0-49) | 0 (0 to 10, 0-70) |
| Maximal knee joint pain experienced during usual care exercise supervised, median (IQR, range), VAS-mm | 21 (0 to 35, 0-100) | 25 (0 to 42, 0-88) | 23 (0 to 40, 0-100) |
| Maximal quadriceps muscle pain experienced during usual care exercise supervised, median (IQR, range), VAS-mm | 0 (0 to 7, 0-80) | 0 (0 to 5, 0-65) | 0 (0 to 6, 0-80) |
| Maximal knee joint pain experienced during BFR-LLST supervised session, median (IQR, range), VAS-mm | 0 (0 to 10, 0-93) | 0 (0 to 16, 0-92) | 0 (0 to 12, 0-93) |
| Maximal quadriceps muscle pain experienced during BFR-LLST supervised session, median (IQR, range), VAS-mm | 52 (15 to 71, 0-100) | 40 (0 to 68, 0-100) | 47 (12 to 70, 0-100) |

Abbreviations: BFR-LLST; Blood flow restriction – low load strength training. SD; Standard Deviation. IQR; Interquartile Range. VAS; Visual Analog Scale.
<sup>a</sup>Total number of exercise (BFR/Usual care exercise supervised (n=15), BFR home (n=30), Usual care exercise home (n=48), BFR total (n=45), Usual care exercise total (n=63)). <sup>b</sup>Two patients (meniscus repair (n=1), cartilage repair (n=1)) lowered their limb occlusion pressure applied due to discomfort; one transient and one permanent. <sup>c</sup>Relative applied 80% LOP: LOP minus 20 mmHg (variability) multiplied with 0.8.

**S7 Table 3.** Change per week in thigh circumference, knee joint and quadriceps pain, perceived exertion and training load during the, on average, 11 weeks of BFR-LLST added to usual care exercise intervention period (15 sessions).

| Outcome measure | Change per week |  |  |  |  |  |
| --- | --- | --- | --- | --- | --- | --- |
|  | Cartilage (n=21) |  | Meniscus (n=21) |  | All (n=42) |  |
|  | mean (95% CI) | p-value | mean (95% CI) | p-value | mean (95% CI) | p-value |
| Thigh circumference - operated leg, cm | 0.05 (-0.01 to 0.11) | <0.099 | 0.12 (0.07 to 0.17) | <0.001 | 0.09 (0.05 to 0.13) | <0.001 |
| Thigh circumference - healthy leg, cm | -0.01 (-0.05 to 0.04) | <0.801 | 0.02 (-0.03 to 0.07) | <0.425 | 0.01 (-0.03 to 0.04) | <0.682 |
| Thigh circumference difference (healthy - operated leg), cm | -0.06 (-0.10 to -0.02) | <0.006 | -0.10 (-0.14 to -0.06) | <0.001 | -0.08 (-0.11 to -0.05) | <0.001 |
| Knee joint pain during BFR-LLST, VAS-mm | -0.3 (-1.1 to 0.5) | <0.407 | -1.1 (-2.0 to -0.3) | <0.010 | -0.7 (-1.3 to -0.1) | <0.017 |
| Quadriceps muscle pain during BFR-LLST, VAS-mm | -0.2 (-1.48 to 1.15) | <0.801 | -0.7 (-1.9 to -0.5) | <0.241 | -0.4 (-1.3 to 0.4) | <0.330 |
| Rating of perceived exertion (8-20-point Borg Scale) during BFR-LLST, points | 0.1 (0.0 to 0.3) | <0.034 | 0.2 (0.1 to 0.3) | <0.001 | 0.2 (0.1 to 0.2) | <0.001 |
| Knee joint pain during usual care exercise, VAS-mm | -0.9 (-2.0 to 0.3) | <0.136 | -0.6 (-1.7 to 0.4) | <0.231 | -0.7 (-1.4 to 0.1) | <0.066 |
| Training load during BFR-LLST, kg | 0.30 (0.22 to 0.38) | <0.001 | 0.26 (0.21 to 0.31) | <0.001 | 0.28 (0.23 to 0.32) | <0.001 |

Abbreviations: CI; Confidence Interval. BFR-LLST; Blood flow restriction-low load strength training. VAS; Visual Analog Scale.

**S7 Table 4.** Number of exercises performed during the group-based usual care exercise supervised program (15 sessions)

| Training modality | Exercise* | Number<br>Mean ± SD (min. to max.) | Total |
| --- | --- | --- | --- |
| Warm-up | Bike | 9 ± 2 (5 to 12) | 385 |
| | Rowing machine | 0 ± 0 (0 to 0) | 0 |
| Warm-up in gym | Slides: Sliding walk | 8 ± 2 (3 to 11) | 347 |
| | Forward lunge | 3 ± 2 (0 to 8) | 121 |
| | Walking lunges with dumbbells | 0 ± 0 (0 to 2) | 8 |
| | Mini Band Side Steps | 8 ± 2 (4 to 12) | 331 |
| Hip abduction strength | The clam w/ elastics | 4 ± 3 (0 to 11) | 176 |
| | Side Plank on Knees with Leg Raises | 4 ± 2 (0 to 8) | 148 |
| | Abduction of the hip with elastic band | 2 ± 2 (0 to 8) | 80 |
| Hamstring strength | "Bridge" | 2 ± 2 (0 to 9) | 75 |
| | Single Leg Glute Bridge, side view | 7 ± 2 (2 to 11) | 301 |
| | Barbell Glute Bridge | 1 ± 1 (0 to 4) | 27 |
| | Romanian Deadlift 2 | 0 ± 1 (0 to 6) | 16 |
| | Single Leg Dumbbell Deadlift | 1 ± 2 (0 to 8) | 42 |
| Hip adductor strength | On your back "squeeze" of ball 1 | 3 ± 2 (0 to 11) | 129 |
| | Side-lying single leg lift 2 | 5 ± 3 (0 to 10) | 218 |
| | Standing inward leg pull w/elastic band 2 | 1 ± 1 (0 to 5) | 25 |
| Calf strength | Elevated Calf Raises | 1 ± 1 (0 to 6) | 58 |
| | Elevated Calf Raises w/ dumbbells | 2 ± 1 (0 to 6) | 69 |
| | Elevated Calf Raises – One-legged | 3 ± 2 (0 to 10) | 134 |
| Knee-extension strength | Cybex: Seated Leg Press 2 | 2 ± 2 (0 to 10) | 66 |
| | Squat 1 | 3 ± 3 (0 to 11) | 123 |
| | Goblet Squat | 0 ± (0 to 1) | 5 |
| | Slides: Back Lunge | 3 ± 2 (0 to 9) | 142 |
| | Slides: Back Lunge w/ dumbbells | 1 ± 2 (0 to 7) | 31 |
| | Bulgarian Split Squat w/barbell | 0 ± 0 (0 to 2) | 2 |
| Balance and Stability | BOSU-ball: balance 1 | 0 ± 1 (0 to 2) | 15 |
| | Balance board: single leg balance 1 | 0 ± 1 (0 to 2) | 13 |
| | BOSU-ball: Balance 3 | 0 ± 0 (0 to 0) | 0 |
| | BOSU-ball: Balance w/ weightball 2 | 0 ± 0 (0 to 0) | 0 |
| Stretching | The Extender | 0 ± 0 (0 to 1) | 1 |
| | Buttocks 2 | 0 ± 0 (0 to 0) | 0 |
| | Hip flexors 1 | 0 ± 1 (0 to 3) | 7 |
| | Stretching of the front thigh 1 | 0 ± 1 (0 to 5) | 17 |
| Other treatments | Otreat (situps, manual mobilization, plank) | 7 ± (0 to 12) | 277 |

\*Exercise names are corresponding to exercise names in S6 Usual care exercise after cartilage and meniscus repair in the knee joint - week 7 postoperatively

**S7 Table 5.** Adverse events during the BFR-LLST added to usual care exercise intervention period

| Adverse events | Cartilage (n=21) | Meniscus (n=21) | Total (n=42) |
| --- | --- | --- | --- |
| Dizziness, events/patients, no | 25/9 | 27/10 | 52/19 |
| Quadriceps muscle pain at rest, events/patients, no | 0/0 | 1/0 | 1/1 |
| Knee joint pain at rest, events/patients, no | 3/2 | 2/2 | 5/4 |
| Numbness (transient), events/patients, no | 1/1 | 1/1 | 2/2 |
| Bruises, events/patients, no, events/patients, no | 2/2 | 2/1 | 4/3 |
| Cardiovascular complaints, events/patients, no | 0 | 0/0 | 0/0 |
| Deep Venous Thrombosis, events/patients, no | 0 | 0/0 | 0/0 |
| Other, events/patients, no<br>(Itching feelings in foot/lower leg, calf tightness, knee pain during exercise, discoloration, discomfort, slight nausea), events/patients, no | 73/19 | 73/19 | 146/38 |

**S7 Table 6.** Average and overall change in knee joint pain, quadriceps muscle pain and perceived exertion from 1^st^ to 4^th^ set within the BFR-LLST session (15 sessions) for all patients (n=42)

| Outcome measure | Change from set 1 |  | Value per set<sup>a</sup> |
| --- | --- | --- | --- |
|  | Mean (95% CI) | p-value | Mean (95% CI) |
| Knee joint pain, VAS-mm |  |  |  |
| Set 1 |  |  | 5.9 (4.8 to 7.0) |
| Set 2 | -0.7 (-2.0 to 0.5) | <0.260 | 5.1 (4.2 to 6.1) |
| Set 3 | -0.6 (-1.9 to 0.6) | <0.328 | 5.1 (4.1 to 6.1) |
| Set 4 | -0.9 (-2.2 to 0.4) | <0.197 | 4.8 (3.7 to 5.9) |
| Type III test |  | <0.224 |  |
| Quadriceps muscle pain, VAS-mm |  |  |  |
| Set 1 |  |  | 11.1 (9.9 to 12.4) |
| Set 2 | 11.3 (9.3 to 13.3) | <0.0001 | 22.4 (20.7 to 24.2) |
| Set 3 | 21.3 (19.2 to 23.3) | <0.0001 | 32.2 (29.9 to 34.5) |
| Set 4 | 30.7 (28.6 to 32.7) | <0.0001 | 40.7 (37.8 to 43.6) |
| Type III test |  | <0.0001 |  |
| Rating of perceived exertion (8-20 point Borg Scale), points |  |  |  |
| Set 1 |  | <0.0001 | 10.8 (10.6 to 11.0) |
| Set 2 | 3.2 (3.0 to 3.4) | <0.0001 | 14.0 (13.8 to 14.1) |
| Set 3 | 5.5 (5.3 to 5.8) | <0.0001 | 16.3 (16.1 to 16.5) |
| Set 4 | 7.8 (7.6 to 8.1) | <0.0001 | 18.5 (18.3 to 18.7) |
| Type III test |  | <0.0001 |  |

BFR-LLST; Blood flow restriction - Low load strength training. CI; Confidence Interval. VAS; Visual Analog Scale. <sup>a</sup>Value per set is an average of a specific set performed during 15 BFR-LLST supervised sessions for all patients (n=42)

**S7 Table 7.**
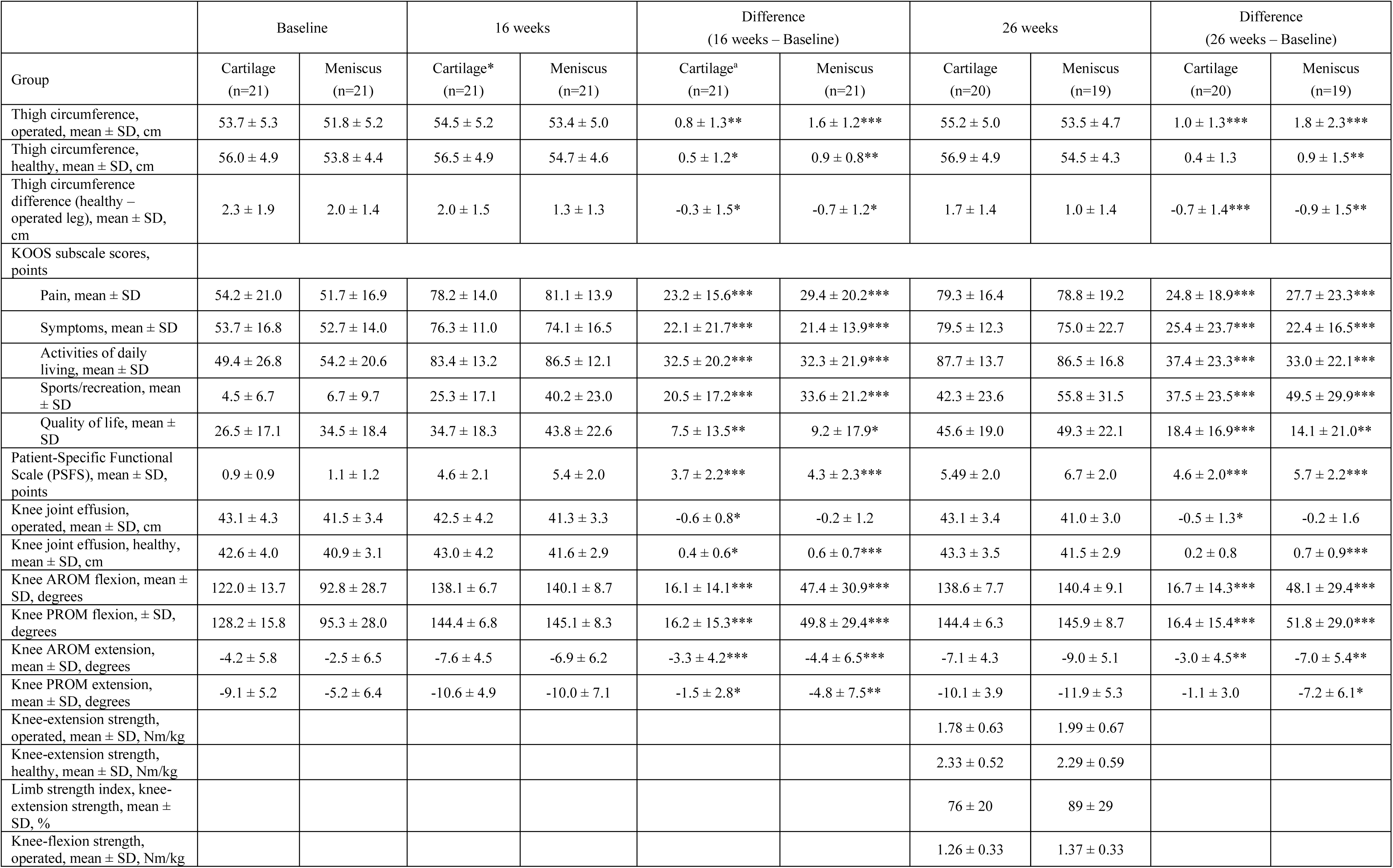
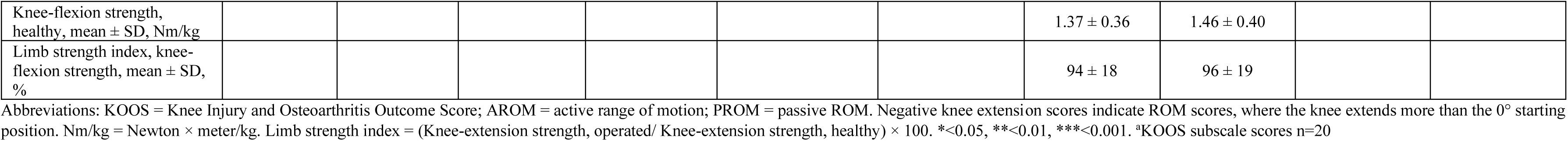
Outcome measures at baseline, 16- and 26-week assessment

|  | Baseline |  | 16 weeks |  | Difference<br>(16 weeks – Baseline) |  | 26 weeks |  | Difference<br>(26 weeks – Baseline) |  |
| --- | --- | --- | --- | --- | --- | --- | --- | --- | --- | --- |
| Group | Cartilage<br>(n=21) | Meniscus<br>(n=21) | Cartilage*<br>(n=21) | Meniscus<br>(n=21) | Cartilage<sup>a</sup><br>(n=21) | Meniscus<br>(n=21) | Cartilage<br>(n=20) | Meniscus<br>(n=19) | Cartilage<br>(n=20) | Meniscus<br>(n=19) |
| Thigh circumference, operated, mean ± SD, cm | 53.7 ± 5.3 | 51.8 ± 5.2 | 54.5 ± 5.2 | 53.4 ± 5.0 | 0.8 ± 1.3** | 1.6 ± 1.2*** | 55.2 ± 5.0 | 53.5 ± 4.7 | 1.0 ± 1.3*** | 1.8 ± 2.3*** |
| Thigh circumference, healthy, mean ± SD, cm | 56.0 ± 4.9 | 53.8 ± 4.4 | 56.5 ± 4.9 | 54.7 ± 4.6 | 0.5 ± 1.2* | 0.9 ± 0.8** | 56.9 ± 4.9 | 54.5 ± 4.3 | 0.4 ± 1.3 | 0.9 ± 1.5** |
| Thigh circumference difference (healthy – operated leg), mean ± SD, cm | 2.3 ± 1.9 | 2.0 ± 1.4 | 2.0 ± 1.5 | 1.3 ± 1.3 | -0.3 ± 1.5* | -0.7 ± 1.2* | 1.7 ± 1.4 | 1.0 ± 1.4 | -0.7 ± 1.4*** | -0.9 ± 1.5** |
| KOOS subscale scores, points |  |  |  |  |  |  |  |  |  |  |
| Pain, mean ± SD | 54.2 ± 21.0 | 51.7 ± 16.9 | 78.2 ± 14.0 | 81.1 ± 13.9 | 23.2 ± 15.6*** | 29.4 ± 20.2*** | 79.3 ± 16.4 | 78.8 ± 19.2 | 24.8 ± 18.9*** | 27.7 ± 23.3*** |
| Symptoms, mean ± SD | 53.7 ± 16.8 | 52.7 ± 14.0 | 76.3 ± 11.0 | 74.1 ± 16.5 | 22.1 ± 21.7*** | 21.4 ± 13.9*** | 79.5 ± 12.3 | 75.0 ± 22.7 | 25.4 ± 23.7*** | 22.4 ± 16.5*** |
| Activities of daily living, mean ± SD | 49.4 ± 26.8 | 54.2 ± 20.6 | 83.4 ± 13.2 | 86.5 ± 12.1 | 32.5 ± 20.2*** | 32.3 ± 21.9*** | 87.7 ± 13.7 | 86.5 ± 16.8 | 37.4 ± 23.3*** | 33.0 ± 22.1*** |
| Sports/recreation, mean ± SD | 4.5 ± 6.7 | 6.7 ± 9.7 | 25.3 ± 17.1 | 40.2 ± 23.0 | 20.5 ± 17.2*** | 33.6 ± 21.2*** | 42.3 ± 23.6 | 55.8 ± 31.5 | 37.5 ± 23.5*** | 49.5 ± 29.9*** |
| Quality of life, mean ± SD | 26.5 ± 17.1 | 34.5 ± 18.4 | 34.7 ± 18.3 | 43.8 ± 22.6 | 7.5 ± 13.5** | 9.2 ± 17.9* | 45.6 ± 19.0 | 49.3 ± 22.1 | 18.4 ± 16.9*** | 14.1 ± 21.0** |
| Patient-Specific Functional Scale (PSFS), mean ± SD, points | 0.9 ± 0.9 | 1.1 ± 1.2 | 4.6 ± 2.1 | 5.4 ± 2.0 | 3.7 ± 2.2*** | 4.3 ± 2.3*** | 5.49 ± 2.0 | 6.7 ± 2.0 | 4.6 ± 2.0*** | 5.7 ± 2.2*** |
| Knee joint effusion, operated, mean ± SD, cm | 43.1 ± 4.3 | 41.5 ± 3.4 | 42.5 ± 4.2 | 41.3 ± 3.3 | -0.6 ± 0.8* | -0.2 ± 1.2 | 43.1 ± 3.4 | 41.0 ± 3.0 | -0.5 ± 1.3* | -0.2 ± 1.6 |
| Knee joint effusion, healthy, mean ± SD, cm | 42.6 ± 4.0 | 40.9 ± 3.1 | 43.0 ± 4.2 | 41.6 ± 2.9 | 0.4 ± 0.6* | 0.6 ± 0.7*** | 43.3 ± 3.5 | 41.5 ± 2.9 | 0.2 ± 0.8 | 0.7 ± 0.9*** |
| Knee AROM flexion, mean ± SD, degrees | 122.0 ± 13.7 | 92.8 ± 28.7 | 138.1 ± 6.7 | 140.1 ± 8.7 | 16.1 ± 14.1*** | 47.4 ± 30.9*** | 138.6 ± 7.7 | 140.4 ± 9.1 | 16.7 ± 14.3*** | 48.1 ± 29.4*** |
| Knee PROM flexion, ± SD, degrees | 128.2 ± 15.8 | 95.3 ± 28.0 | 144.4 ± 6.8 | 145.1 ± 8.3 | 16.2 ± 15.3*** | 49.8 ± 29.4*** | 144.4 ± 6.3 | 145.9 ± 8.7 | 16.4 ± 15.4*** | 51.8 ± 29.0*** |
| Knee AROM extension, mean ± SD, degrees | -4.2 ± 5.8 | -2.5 ± 6.5 | -7.6 ± 4.5 | -6.9 ± 6.2 | -3.3 ± 4.2*** | -4.4 ± 6.5*** | -7.1 ± 4.3 | -9.0 ± 5.1 | -3.0 ± 4.5** | -7.0 ± 5.4** |
| Knee PROM extension, mean ± SD, degrees | -9.1 ± 5.2 | -5.2 ± 6.4 | -10.6 ± 4.9 | -10.0 ± 7.1 | -1.5 ± 2.8* | -4.8 ± 7.5** | -10.1 ± 3.9 | -11.9 ± 5.3 | -1.1 ± 3.0 | -7.2 ± 6.1* |
| Knee-extension strength, operated, mean ± SD, Nm/kg |  |  |  |  |  |  | 1.78 ± 0.63 | 1.99 ± 0.67 |  |  |
| Knee-extension strength, healthy, mean ± SD, Nm/kg |  |  |  |  |  |  | 2.33 ± 0.52 | 2.29 ± 0.59 |  |  |
| Limb strength index, knee-extension strength, mean ± SD, % |  |  |  |  |  |  | 76 ± 20 | 89 ± 29 |  |  |
| Knee-flexion strength, operated, mean ± SD, Nm/kg |  |  |  |  |  |  | 1.26 ± 0.33 | 1.37 ± 0.33 |  |  |
| Knee-flexion strength, healthy, mean ± SD, Nm/kg | | | | | | | 1.37 ± 0.36 | 1.46 ± 0.40 | | |
| Limb strength index, knee-flexion strength, mean ± SD, % | | | | | | | 94 ± 18 | 96 ± 19 | | |

Abbreviations: KOOS = Knee Injury and Osteoarthritis Outcome Score; AROM = active range of motion; PROM = passive ROM. Negative knee extension scores indicate ROM scores, where the knee extends more than the 0° starting position. Nm/kg = Newton × meter/kg. Limb strength index = (Knee-extension strength, operated/ Knee-extension strength, healthy) × 100. \* $<0.05$ , \*\* $<0.01$ , \*\*\* $<0.001$ . <sup>a</sup>KOOS subscale scores n=20

## Notes

### Competing Interest Statement

The authors have declared no competing interest.

### Clinical Trial

Clinical.Trials.gov (NCT03371901)

### Author Declarations

The Committees on Biomedical Research Ethics for the Capital Region of Denmark approved the study (Protocol nr. 17010473)

